# Impact of a school-based water and hygiene intervention on child health and school attendance in Addis Ababa, Ethiopia: a cluster-randomised controlled trial

**DOI:** 10.1101/2024.04.08.24305021

**Authors:** Sarah Bick, Alem Ezezew, Charles Opondo, Baptiste Leurent, Wossen Argaw, Erin C Hunter, Oliver Cumming, Elizabeth Allen, Robert Dreibelbis

## Abstract

**Background:** Water, sanitation and hygiene (WASH) interventions in schools may improve the health and school attendance of schoolchildren, particularly among post-menarcheal girls, but existing evidence is mixed. We examined the impact of an urban WASH in schools programme (Project WISE) on child health and attendance.

**Methods:** The WISE cluster-randomised trial, conducted in 60 public primary schools in Addis Ababa, Ethiopia over one academic year, enrolled 2–4 randomly selected classes per school (approximately 100 pupils) from grades 2–8 (aged 7–16) in an ’open cohort’. Schools were assigned 1:1 by stratified randomisation to receive the intervention during the 2021/22 academic year or the 2022/23 academic year (waitlist control). Masking was not possible. The intervention included improvements to drinking water storage, filtration and access, alongside handwashing stations and behaviour change promotion. Planned improvements to sanitation facilities were not realised. At four unannounced classroom visits between March and June 2022 (post-intervention, approximately every four weeks), enumerators recorded primary outcomes of roll-call absence, and pupil-reported respiratory illness and diarrhoea in the past seven days among pupils present. Analysis was by intention-to-treat. This study is registered with ClinicalTrials.gov, number NCT05024890.

**Findings:** Of 83 eligible schools, 60 were randomly selected and assigned. In total, 6229 eligible pupils were enrolled (median per school 101·5; IQR 94–112), with 5987 enrolled at study initiation (23rd November–22nd December 2021) and the remaining 242 during follow-up. Data were available on roll-call absence for 6166 pupils (99·0%), and on pupil-reported illness for 6145 pupils (98·6%). We observed a 16% relative reduction in the odds of pupil-reported respiratory illness in the past seven days during follow-up in intervention schools vs. control schools (aOR 0·84; 95% CI 0·71– 1·00; p=0·046). No effect was observed on pupil-reported diarrhoea in the past seven days (aOR 1·15; 95% CI 0·84–1·59; p=0·39) nor roll-call absence (aOR 1·07; 95% 0·83–1·38; p=0·59). There was a small increase in menstrual care self-efficacy (aMD 3·32 on 0–100 scale; 95% CI 0·05–6·59), and no effects on the other health, wellbeing and absence secondary outcomes.

**Interpretation:** This large-scale intervention to improve WASH conditions in schools across a large city had a borderline impact on respiratory illness among schoolchildren but no effect on diarrhoeal disease nor pupil absence. Future research should establish the relationships between WASH-related illness and absence and other downstream educational outcomes.

**Funding:** Children’s Investment Fund Foundation.

***Panel:* Research in context:** *Evidence before this study:* Prior to this study, there were several systematic reviews on water, sanitation and hygiene (WASH) in schools, none of which used meta-analysis methods due to heterogeneity in intervention components and outcome measures. In the most comprehensive review in 2019, McMichael reported mixed evidence for the effectiveness of WASH in schools in low-income countries across health and educational outcomes, and randomised and non-randomised studies. Prior to starting this trial, there were two randomised trials conducted exclusively in urban schools in a low- or middle-income country: a handwashing trial in Cairo, Egypt examining absence due to influenza, and a trial of hand sanitizer and respiratory hygiene in Dhaka, Bangladesh for reducing influenza-like illness and laboratory-confirmed influenza. During this study, another randomised trial in Manila, Philippines was published, with different outcomes (malnutrition and dehydration). The effectiveness of combined water, sanitation and hygiene in urban schools on respiratory illness, diarrhoea and overall absence was not known, and some previous evaluations have used school records alone to track attendance. Previous studies evaluating WASH in schools interventions have suggested that multi-component interventions may be more effective, and that specific effects on girls’ absence might be observed with provision of a safe, private space to change menstrual materials.

*Added value of this study:* This study provides rigorous experimental evidence on the effectiveness of an urban school-based water and hygiene intervention in reducing pupil-reported respiratory illness among schoolchildren, during the COVID-19 pandemic. There was no evidence of effects on pupil-reported diarrhoea or absence, nor gender-specific effects on absence. We highlight the value of unannounced visits for absence tracking with comparison to pupil-reported absence, and the need to distinguish seasonal and pandemic illness in future trials.

*Implications of all the available evidence:* Our results are consistent with the mixed impacts on health and absence found in previous WASH in schools evaluations. The lack of detected effects on diarrhoea, attendance and secondary outcomes related to wellbeing and menstrual health should be considered in light of the absence of sanitation infrastructure improvements, which were not delivered until after trial completion, which may have influenced risk of diarrhoeal disease. Nonetheless, school absence is multi-factorial, and these findings temper expectations that absence can be impacted by reductions in one domain of illness and not the many other important drivers linked to poverty and gender, and few programmes are likely to be able to obtain a more ambitious infrastructure and behaviour change programme at the scale of the one included in this trial, which is currently being replicated in other cities across Ethiopia.

## Introduction

School-aged children in low- and middle-income countries (LMICs) are particularly susceptible to water, sanitation and hygiene (WASH)-related morbidities including gastrointestinal^1^ and respiratory infections,^2^ often due to frequent social mixing.^3^ In low-resource settings, these health risks are particularly associated with absence from school, lower test scores and dropout,^4,5^ with implications on downstream social, occupational and health outcomes.^6^ WASH interventions in schools are often expected to have gendered impacts: while reasons for absence and dropout are varied,^5^ inadequate WASH conditions in schools may present barriers to attendance, including through lack of hygienic menstrual materials, disposal facilities and privacy leaving girls with limited options for menstrual hygiene management (MHM), and impacting educational progression.^7^ Pupils’ academic performance may also be affected by dehydration where there is inadequate water supply.^8^

Although WASH in schools interventions have been hypothesised to improve children’s health and attendance outcomes, evidence of their impact has been mixed. A systematic review of varied WASH in schools intervention studies in low-income countries, including provision of water for drinking and handwashing, water quality, sanitation, and hygiene promotion,^9^ found significant reductions in pupil-reported diarrhoeal disease between 29% and 50%, and reduced incidence of respiratory illness. Other studies, however, found no significant impacts or saw positive impacts for only select disease outcomes.^10,11^ Mixed health effects are observed in other randomised trials in urban settings.^12–14^ Impact on absence is similarly ambiguous: WASH improvements have been shown to reduce absence,^9,15^ but only one^10^ randomised controlled trial reports significantly lower overall absence rates. Some studies observed specific impacts on girls’ absence alone,^16^ or on absence due to diarrhoea.^17^

Access to safe WASH facilities in school environments is included under Sustainable Development Goal (SDG) 6^18^ as essential in ensuring dignity and equity, and promoting women’s equality and empowerment. To achieve access to safe WASH, interventions must ensure sustained management of water and sanitation services over time^11^, including consistent availability of soap and water for practicing handwashing.^19^ Several publications highlight that combined WASH interventions versus single interventions – such as handwashing alone – may be necessary to transform school environments to the extent that the risk of illness and absence is reduced.^9^ However there is limited robust evidence for the effectiveness of combined interventions delivered at-scale in urban settings.

The aim of this trial was to evaluate the effectiveness of a large-scale urban WASH in schools intervention, including water and sanitation infrastructure, behaviour change promotion and targeted MHM services, in schools in Addis Ababa, Ethiopia. ‘Project WISE (WASH in Schools for Everyone)’, implemented by US-based NGO *Splash* in Addis Ababa public schools, is being delivered to pre-defined groups of schools on an annual basis. We hypothesised that the intervention would improve child health and school attendance, with greater impacts among post-menarcheal girls, and used unannounced attendance checks to avoid bias commonly associated with absence measurement.^20^

## Methods

### Study design

The WISE evaluation was a parallel two-arm school-based cluster-randomised controlled trial, with 60 public primary schools in Addis Ababa, Ethiopia constituting the study clusters. We used a cluster-randomised design because the intervention evaluated was delivered at the school level and comprised changes to the whole school environment. Seventeen additional kindergarten schools were enrolled as part of a sub-study estimating the impact of Project WISE on kindergarten pupils, to be reported in a separate publication.

The trial was conducted over the course of one Ethiopian academic year (November 2021 to July 2022; schools were open from September to July) and followed an ‘open cohort’ design to minimise participant attrition. Between two and four sentinel classrooms of pupils were randomly selected for follow-up during the year, with pupils who joined the class late, left the class, or were absent from the first visit contributing data to analysis. Follow-up consisted of four unannounced visits to sentinel classrooms post-intervention (approximately every four weeks), concurrently in intervention and control arms.

The study protocol (Supplementary File A) was approved by the London School of Hygiene & Tropical Medicine Research Ethics Committee (reference 17761), and the National Research Ethics Review Committee of Ethiopia (NRERC; reference A/A/H/10H02/227) prior to commencement of study activities. Under direction of the study investigators, Holster International Research and Development Consultancy was responsible for data collection.

### Participants

The study population comprised primary school pupils aged 7–16 years attending schools due to receive the WISE intervention in either the 2021/2022 or 2022/2023 academic years. In order to have sufficient pupils in the eligible age range, we excluded schools without pupils in grades 2–8. We also excluded schools that received a WASH intervention in the three years prior to study activities and schools that provided education to vulnerable populations only. Sixty schools meeting these criteria were randomly selected for participation.

Within each participating school, between two and four classes were selected from grades 2–8. We obtained enrolment data (number of pupils and classes) for all grades in the school and estimated mean class size for each grade. In order to have sufficient older pupils for age-specific secondary outcome measures, we followed a stratified selection process, selecting one class each from grades 2–5 and grades 6–8, then continued alternating from younger grades and older grades until there were estimated >100 pupils.

All eligible schools had consented to the receipt of the Project WISE intervention. Once the random allocation had been determined, formal consent for participation in the trial was sought from school principals *in loco parentis,* on behalf of all pupils in the school. School principals received guidance for communicating to parents, and parental information sheets and opt-out forms were distributed to all pupils in the sentinel classes at least one week before pupil enrolment; additional information sheets were provided in case of unexpected variation in class sizes and enrolment of additional pupils throughout the year. Pupils were excluded from data collection if their parent or guardian returned the opt-out form at any point and were required to give oral assent before each data collection activity.

Pupils were included in data collection regardless of age in order to minimise risk of social exclusion in the classroom if particular pupils were excluded, but only pupils aged 7–16 at enrolment were included in analysis. Some data collection activities were restricted to subgroups of pupils by age and gender.

### Randomisation and masking

Randomisation was conducted in July 2021, using a random number generator in Stata version 18·0 (StataCorp, College Station, TX, USA). From the list of 143 schools due to receive the intervention, we excluded ineligible schools (as described above), and then randomly selected 60 schools of 83 eligible schools for participation. Randomisation was stratified by school size (< or ≥1200 pupils) and presence of a kindergarten (to facilitate the kindergarten sub-study). Within strata the schools were randomly ranked, and the first half of the schools assigned to the intervention (implementation during the 2021/2022 academic year). The remaining schools were assigned to the waitlist control arm, to receive the intervention in the 2022/2023 academic year, after study completion. Investigators performing the randomisation had no prior knowledge of any of the study schools. Due to the visible, prominent nature of the intervention, masking of school administrators, participants, or those delivering the intervention was not possible. Outcome assessors were not informed of treatment assignment, but might have inferred it, for example, from the distinctive WASH infrastructure components.

### Procedures

The Project WISE intervention combined infrastructure and behaviour change promotion activities, so that handwashing and drinking exclusively from filtered water become normative behaviours, and girls are able to manage menses at school. Intervention design was informed by the behaviour-centred design approach^21^ to alter behaviour through environmental cues, along with activity-based curricula, and pupil and teacher motivators. The intervention was delivered at the school level, so all children attending school were exposed to the intervention regardless of trial participation.

Infrastructure components included correcting water storage capacity deficits through water storage tanks; water filtration systems for drinking water; and durable plastic drinking water and handwashing stations with specific features differentiating their use and installed with taps to meet sufficient tap-to-pupil ratios. Further details and images are provided in Supplementary File B. The intervention also includes provision of new or rehabilitated toilet facilities to meet standards, however this component is managed by the Addis Ababa Education Bureau on a separate timeline and was not delivered to intervention schools until after the evaluation period.

Splash staff conducted a site engagement meeting with school administration and worked with the school to organise a family ‘soap drive’ and ‘menstrual pad drive’ during school registration, whereby families of pupils are encouraged to donate hygiene products to the school to ensure availability of products throughout the year. Two ‘focal teachers’ per school were trained to promote the WASH programme and organise a 20 to 30-pupil ‘hygiene club’ at each school. The one-day training for hygiene focal teachers covered safe water and water conservation, handwashing, personal hygiene, sanitation, and hygiene clubs and action planning. Two additional female focal teachers and one male focal teacher were trained to organise a ‘gender club’ focussed on MHM, which took place over two days with 20–30 girls and 20–30 boys trained on MHM. The gender club focal teachers were trained in puberty, menstrual health and discussing sensitive topics. Focal teachers then organised parent-teacher association orientation and delivered information on MHM to parents. School janitors, maintenance staff, and food handlers also received training on hygiene and operation and maintenance of infrastructure.

Splash staff also supported focal teachers in training the hygiene club members to influence their peers through monitoring handwashing during breaks, ensuring soap availability at handwashing stations, delivering hygiene messaging during school announcements, and assisting in planning event days promoting hygiene school-wide. Members held monthly meetings to track progress and bring issues and requests to school leadership. Within the menstrual health programme, all children aged 10 years and older received an education session on puberty and menstruation, including a Q&A session and product demonstration for girls, and a puberty workshop for boys. Peer mentoring of younger girls by older girls took place over four sessions. Menstrual health event days were also organised.

Interventions included behavioural ‘nudges’^22^ to subtly guide pupils towards the intended behaviours, such as mirrors and posters at handwashing stations, and brightly coloured vests for hygiene club members to wear during handwashing monitoring.

School engagement and training began in November 2021, and all infrastructure components were installed (excluding sanitation infrastructure) and core training modules delivered by January 2022 in all 30 intervention schools. School and pupil enrolment activities took place concurrent with intervention delivery (November to December 2021); therefore, outcomes assessed at enrolment were not included in the primary impact assessment.

Between May and July 2021, data collection tools and methods for school and pupil enrolment and routine follow-up surveys were piloted in five randomly selected schools not included in the primary impact evaluation where Splash implemented the WISE intervention in 2020 / 2021. During piloting, in-country data collection partners followed full study procedures outlined below, with one minor variation: follow-up of sentinel classrooms occurred three weeks after enrolment and only one round of follow-up occurred. The study pilot was used to assess the logistics of field data collection, verify assumptions made in sample size calculations, and test and adapt MHM scales. Minor adjustments to class selection procedures were made as a result.

Data collection activities were completed in one day per school. Following school enrolment and selection of sentinel classes, a team of trained enumerators visited the classrooms and conducted a detailed enrolment survey using tablets, one-to-one with assenting pupils (approximately 15 minutes), including demographic information and household WASH access, self-reported number of full- and partial-days absent in the past week, causes of absence, and symptoms of infectious disease over the preceding two and seven days. These surveys were used to create a digital roster of pupils in sentinel classes, which was automatically updated as new pupils were enrolled or left the class during the academic year. A pupil identification number was assigned internally to all pupils on the roster to anonymously link their data across surveys.

Between March and June 2022, enumerators conducted four unannounced follow-up visits to sentinel classes in each school. At the first three follow-ups, enumerators took attendance using the digital rosters and conducted a brief survey with each pupil present (< five minutes) collecting data only on self-reported absence, causes of absence, diarrhoea and respiratory illness in the past week. Pupils absent from the initial enrolment survey completed the enrolment survey at the first follow-up visit they were present for, and were retrospectively marked as absent from all previous visits conducted while they had been enrolled at the school. If a pupil was absent from two consecutive follow-ups, enumerators were automatically prompted to ask teachers if the pupil had dropped out of school, and, if they had, to note the date of dropout and reason for dropout if known.

At the final follow-up, attendance was taken and all outcomes were assessed, including wellbeing and menstrual health outcomes. Pupils meeting inclusion criteria for the Strengths and Difficulties Questionnaire^23^ (aged 11 and above), and the menstrual health measures (post-menarcheal girls aged ten and above) were identified and given their pupil identification number to link their records with these self-completed paper-based questionnaires. Age or date of birth (if known) and gender were self-reported by pupils at enrolment. At the final follow-up, age and date of birth were double-checked to ensure accuracy and updated. Age at enrolment was calculated based on the updated records.

### Outcomes

All outcomes were measured at the individual participant level. The primary health outcomes were pupil-reported diarrhoea (defined as occurrence of at least three loose stools in a 24-hour period) and pupil-reported respiratory illness (defined as occurrence of cough, sneezing or rhinorrhoea) in the past seven days. Both were recorded at each follow-up visit as dichotomous variables. The primary absence outcome was roll-call absence, recorded at each follow up as a dichotomous variables.

Secondary outcomes were pupil-reported absence (number of full days reported absent out of number of days of reporting in the past week); pupil-reported diarrhoea and pupil-reported respiratory illness in the past two days; Strengths and Difficulties Questionnaire (SDQ-25)^23^ total difficulties score, a widely used measure of pupil behavioural and mental health challenges designed for use among school-aged children that has been used in a number of Sub-Saharan African countries,^24^ measured among children aged 11–16 at final follow-up; Self-efficacy in Addressing Menstrual Needs Scale (SAMNS-26)^25^ total score, a measure of girls’ confidence in addressing their menstrual needs; and Menstrual Practice Needs Scale (MPNS-36)^26^ total score, a measure of how well current menstrual practices are perceived to meet the girls’ needs, with SAMNS-26 and MPNS-36 both measured among post-menarcheal girls aged 10–16 at final follow-up.

Other outcomes were absence due to illness, diarrhoea, and respiratory illness; seven- and two-day occurrence of earache (negative control for illness outcomes, as earache is not feasibly affected by the intervention); child subjective wellbeing assessed through a smiley faces visual analogue (1–5 scale, with 5 being the best mood possible and 1 the worst); Sanitation-related Quality of Life (SanQoL-5) applying attribute weights from a study in Ethiopia [preprint];^27^ change in gender parity in school enrolment over the academic year using the adjusted gender parity index;^28^ and SAMNS-26 and MPNS-36 sub-scales.

### Statistical analysis

Sample size calculations were based on estimating the mean difference between arms in pupil-level proportions of illness or absence across the follow-ups. Assuming the mean pupil-level proportions of follow-ups reporting diarrhoea in the control group was 0·08 (SD 0·05), a two-sided type I error (α) of 0·05, and intracluster correlation coefficient (ICC) of 0·15 (conservatively; ICC estimates for pupil-reported illness outcomes in our pilot study ranged from 0·07–0·10), we estimated 50 schools (25 per arm) with 100 children per school was sufficient to detect a reduction in the mean proportion of follow-ups with diarrhoea of 0·016 (standardised effect size 0·32). This standardised effect size equates to a reduction in mean proportion of follow-ups reporting respiratory infection of 0·084 and a reduction of 0·022 in the mean proportion of follow-ups absent as assessed through roll call at each follow up, based on SD estimates from pilot data. Schools were oversampled to account for cluster attrition; we randomised 60 schools to meet the sample size of 50 schools with 17% attrition.

The statistical analysis plan was pre-registered on 1st March 2023^29^ before allocation was revealed. Analyses were done by intention-to-treat. Characteristics of the children and schools at enrolment were summarised by treatment arm. Statistical analyses of the outcomes were conducted at the individual level with mixed effects regression models, using logistic (for pupil-reported illness, roll-call absence and causes of absence outcomes), binomial (pupil-reported absence i.e. number of days reported absent with offset of number of days of reporting), linear (SDQ-25, SAMNS-26, MPNS-36, SanQoL-5 and gender parity in enrolment), and ordered logistic (subjective wellbeing) regression models as appropriate. We additionally carried out a confirmatory analysis based on the mean proportion of episodes per pupil. Analyses included a random effect for school and analyses based on repeated measures included an additional random effect for pupil-level clustering, and assumed a constant treatment effect across time-points. Primary estimates of effectiveness were calculated using a basic model adjusting for stratification factors alone: school size and presence of kindergarten classes. Further adjustments in secondary analyses were made for school grade and gender, and a fully adjusted model also adjusted for school location by sub-city, and time-point in analyses of repeated measures. Interaction tests were used to examine the differential effect of the intervention by gender, and across time-points on the three primary outcomes. We examined factors associated with missing outcome data (due to absence) at the final follow-up, and conducted exploratory sensitivity analyses of primary outcomes adjusting for factors associated with missingness. Sensitivity analysis including all pupils enrolled in the sentinel classes regardless of age was also performed. We used Stata version 18·0 (StataCorp, College Station, TX, USA) for all analyses.

This trial is registered with ClinicalTrials.gov, number NCT05024890.

## Results

Of the 143 schools due to receive the intervention in 2021/2022 or 2022/2023, 83 were eligible (Figure 1) for the trial. None of the 60 schools that were randomly selected and consented to participation during school enrolment (2nd to 22nd November 2021) withdrew from the study. In total, 6,455 pupils were enrolled at any point in the trial, and 6,229 were later determined to be eligible by age. Of those eligible, 5,987 were enrolled during pupil enrolment (23rd November to 22nd December 2021) and the remaining 242 were enrolled during follow-up, due to absence at enrolment (231) or joining the class midway through the academic year (11). Eighty-two pupils left classes (dropped out of school) during the study, most commonly due to transferring school or leaving the area. The number of pupils contributing data to outcome assessments is shown for each time-point in figure 1 and each analysis table; a detailed summary of observations for each outcome is found in Supplementary Table 1. For roll-call attendance, 6,166 (99·0%) pupils were registered in the sentinel classes (whether or not they were present) during follow-up, i.e. had not dropped out before the first follow-up. For pupil-reported repeated measures, 6,145 (98·6%) were present for at least one follow-up, balanced between study arms.

**Figure 1.**
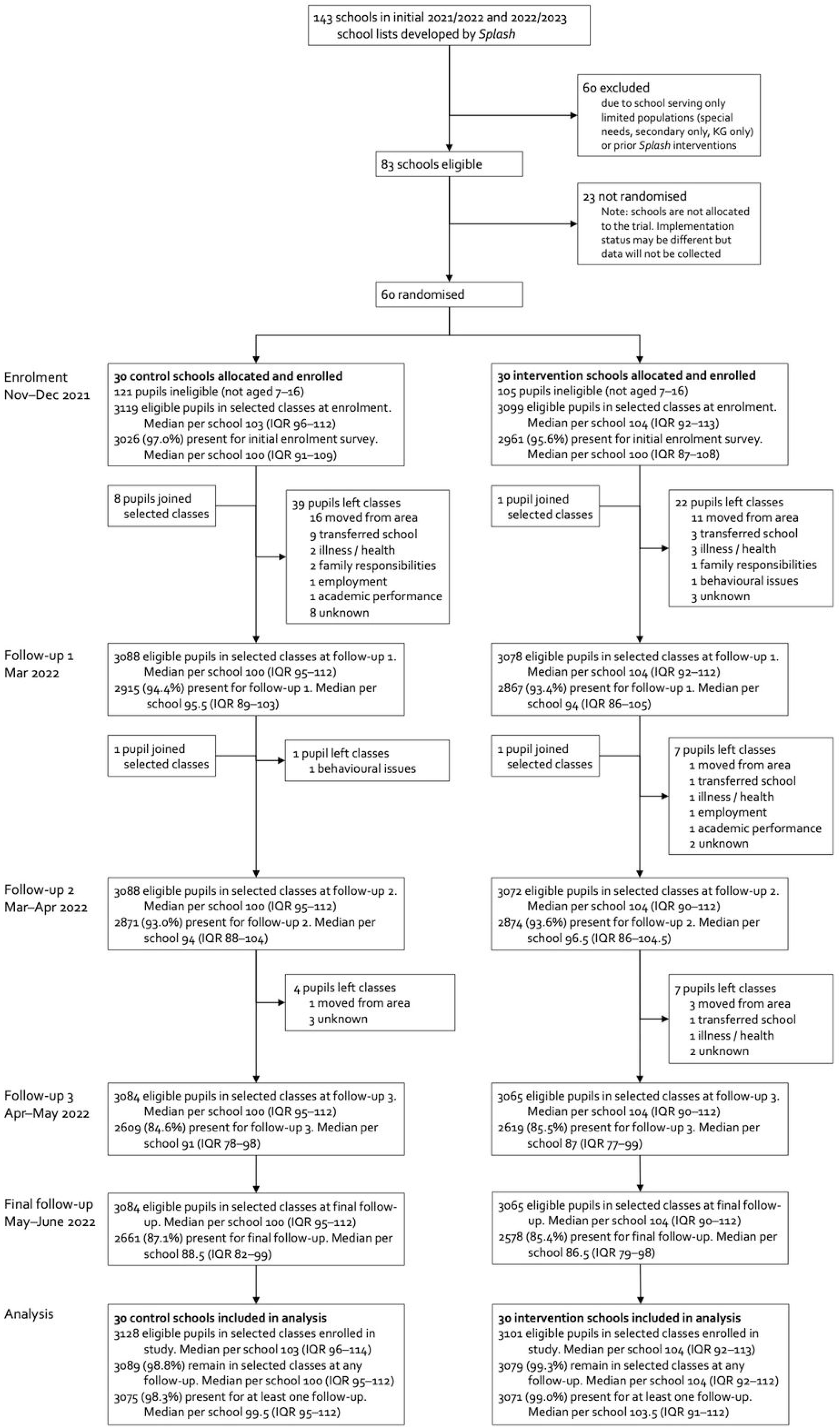
Trial profile.

Of all eligible children enrolled, 52·6% were girls, the mean age was 12·1 years (SD 2·5), few (27%) had at least a basic sanitation service at home, but the majority (63%) reported their household having at least a basic water supply (Table 1). Pupils missing at final follow-up were similar to those present, except earning money for the household was associated with missingness (data not shown). There was a slight difference in school size across arms; other characteristics were balanced. Of the girls aged ten and above at the final follow-up, 48·5% (588/1212) in control schools and 45·3% (581/1063) in intervention schools had reached menarche, with median reported age at menarche 13 in both arms.

**Table 1.** Characteristics of participants and schools at enrolment.

|  | *Control* | *Intervention* |
| --- | --- | --- |
| Number of pupils | 3128 | 3101 |
| Age in years, mean (SD) | 12.2 (2.5) | 12.0 (2.5) |
| Female gender | 1674 (53.5%) | 1603 (51.7%) |
| Time taken to travel to school, median (IQR) | 15 (10, 30) | 15 (10, 30) |
| Household responsibilities |  |  |
| Collecting water | 1666 (53.4%) | 1741 (56.2%) |
| Earning money for household | 75 (2.4%) | 72 (2.3%) |
| Childcare | 973 (31.1%) | 945 (30.5%) |
| Household members, median (IQR) | 5 (4, 6) | 5 (4, 6) |
| At least basic household water supply* | 1912 (61.1%) | 1993 (64.3%) |
| At least basic household sanitation† | 857 (27.4%) | 815 (26.3%) |
| Number of schools | 30 | 30 |
| Sub-city |  |  |
| Addis Ketema | 3 (10.0%) | 2 (6.7%) |
| Akaky Kality | 2 (6.7%) | 6 (20.0%) |
| Arada | 3 (10.0%) | 4 (13.3%) |
| Bole | 2 (6.7%) | 4 (13.3%) |
| Gulelle | 3 (10.0%) | 3 (10.0%) |
| Kirkos | 5 (16.7%) | 2 (6.7%) |
| Kolfe Keraniyo | 5 (16.7%) | 3 (10.0%) |
| Lemi Kura | 1 (3.3%) | 1 (3.3%) |
| Lideta | 1 (3.3%) | 3 (10.0%) |
| Nifas Silk Lafto | 5 (16.7%) | 2 (6.7%) |
| Number of grades, median (IQR) | 8 (8, 8) | 8 (8, 8) |
| Number of pupils, median (IQR) | 1112.5 (400, 1527) | 935.5 (618, 1617) |
| Gender parity in enrolment,‡ mean (SD) | 1.09 (0.11) | 1.10 (0.10) |
| Number of disabled pupils, median (IQR) | 33 (14, 75) | 34 (18, 68) |
| Mean class size, mean (SD) | 48.8 (12.7) | 46.3 (9.3) |
| Number of classes enrolled in the study |  |  |
| 2 | 22 (73.3%) | 19 (63.3%) |
| 3 | 5 (16.7%) | 10 (33.3%) |
| 4 | 3 (10.0%) | 1 (3.3%) |

\*Defined as an improved facility not shared with other households. †Defined as drinking water from an improved source, with collection time not more than 30 minutes roundtrip. ‡Adjusted gender parity index.<sup>28</sup>

The mean proportion of follow-ups where pupils reported diarrhoea in the past seven days (co-primary health outcome) was 0·073 in control schools and 0·083 in intervention schools (Table 2), with no significant difference between study arms in the primary analysis adjusting for clustering and stratification factors (aOR 1·15; 95% CI 0·84 to 1·59; p=0·39). The mean proportion of follow-ups reporting respiratory illness in the past seven days (co-primary health outcome) was 0·278 in control schools and 0·248 in intervention schools, corresponding to a 16% relative reduction in the odds of pupil-reported respiratory illness in the past seven days during follow-up in the primary analysis (aOR 0·84; 95% CI 0·71 to 1·00; p=0·046). Mean proportion of follow-ups absent from roll-call was similar between arms (0·103 in control schools vs. 0·106 in intervention schools), with no significant difference in odds of absence (aOR 1·07; 95% 0·83 to 1·38; p=0·59) in the primary analysis.

**Table 2.** Intervention effects on primary and secondary outcomes.

|  | Control |  | Intervention |  | Intervention effect |  |  |
| --- | --- | --- | --- | --- | --- | --- | --- |
|  | Pupils | Mean prop. of follow-ups with illness (SD) | Pupils | Mean prop. of follow-ups with illness (SD) | aOR* (95% CI) | p-value | ICC |
| Pupil-reported diarrhoea in past 7 days† | 3075 | 0.073 (0.151) | 3069 | 0.083 (0.160) | 1.15 (0.83, 1.59) | 0.39 | 0.088 |
| Pupil-reported diarrhoea in past 2 days | 3075 | 0.042 (0.112) | 3070 | 0.050 (0.127) | 1.22 (0.82, 1.83) | 0.32 | 0.13 |
| Pupil-reported respiratory illness in past 7 days† | 3075 | 0.276 (0.269) | 3070 | 0.248 (0.257) | 0.84 (0.71, 1.00) | 0.046 | 0.025 |
| Pupil-reported respiratory illness in past 2 days | 3075 | 0.187 (0.230) | 3070 | 0.171 (0.219) | 0.88 (0.73, 1.06) | 0.18 | 0.029 |
|  | Pupils | Mean prop. of follow-ups absent (SD) | Pupils | Mean prop. of follow-ups absent (SD) | aOR* (95% CI) | p-value | ICC |
| Roll-call absence† | 3088 | 0.103 (0.171) | 3078 | 0.106 (0.171) | 1.07 (0.83, 1.38) | 0.59 | 0.051 |
|  | Pupils | Mean prop. of school days reported absent (SD) | Pupils | Mean prop. of school days reported absent (SD) | aOR* (95% CI) | p-value | ICC |
| Pupil-reported full-day absence in past week | 3075 | 0.056 (0.090) | 3070 | 0.055 (0.082) | 1.01 (0.75, 1.36) | 0.93 | 0.067 |
|  | Pupils | Mean score (SD) | Pupils | Mean score (SD) | aMD (95% CI) | p-value | ICC |
| SDQ-15 total difficulties score (0–40) | 1829 | 10.1 (6.0) | 1675 | 10.0 (6.0) | 0.03 (-0.62, 0.68) | 0.94 | 0.028 |
| SAMNS-26 total score (0–100) | 545 | 69.1 (17.9) | 438 | 72.3 (18.6) | 3.32 (0.05, 6.59) | 0.046 | 0.060 |
| MPNS-36 total score (0–3) | 530 | 1.92 (0.39) | 406 | 1.91 (0.39) | -0.01 (-0.08, 0.06) | 0.81 | 0.051 |

*Note:* Analyses include all eligible children with outcome data at the relevant follow-up. For repeated measures, we calculated a proportion of available follow-ups with illness / absent for each participant, and the mean and SD of these proportions across participants are shown. Analyses were adjusted for stratification variables (school size < or ≥1200 pupils, and presence of a kindergarten), with further adjusted models reported in Supplementary Table 2. All analyses included a random effect for school, and analyses based on repeated measures included an additional random effect for pupil-level clustering and assumed a constant treatment effect across time-points. \*For dichotomous outcomes, the estimated differences in means are reported in Supplementary Table 3. †Primary outcomes.

Models with further covariate adjustments produced similar results (Supplementary Table 2), as did analysis based on mean differences in pupil-level proportions (Supplementary Table 3).

Among the secondary outcomes, effects on pupil-reported diarrhoea and respiratory illness in the past two days were similar in direction to the respective seven-day outcomes but with no evidence of differences between arms (Table 2). Pupils reported absence at a much lower rate than roll-call absence (mean proportion of school days reported absent was 0·056 in control schools and 0·055 in intervention schools), with no evidence of a difference between arms. We observed a small increase in SAMNS-26 total score in the intervention arm vs. control of three points on a 0–100 scale (mean difference 3·32; 95% CI 0·05 to 6·59; p=0·046). There was no evidence of differences in either MPNS-36 total score or SDQ-25 total difficulties score between arms.

We observed no evidence of effects on other outcomes, including causes of absence, pupil-reported earache (negative control for pupil-reported illness), subjected wellbeing measured through a smiley faces visual analogue scale, menstrual health sub-scales, and gender parity in enrolment (Supplementary Table 4), with the exception of past-week absence due to diarrhoea (aOR 0·59; 95% CI 0·37 to 0·93; p=0·024), which was very rarely reported (mean proportion of follow-ups of 0·008 in controls schools vs. 0·005 in intervention schools).

There was some evidence of effect modification by gender (p=0·021) for pupil-reported respiratory illness in the past seven days, with a greater intervention effect observed in boys (Figure 2). Findings for pupil-reported diarrhoea and roll-call absence were consistent across genders (p-value for interaction 0·96 and 0·54, respectively). There was no evidence of group-time interaction for pupil-reported respiratory illness (p=0·31) or diarrhoea (p=0·67), or roll-call absence (p=0·89).

**Figure 2.**
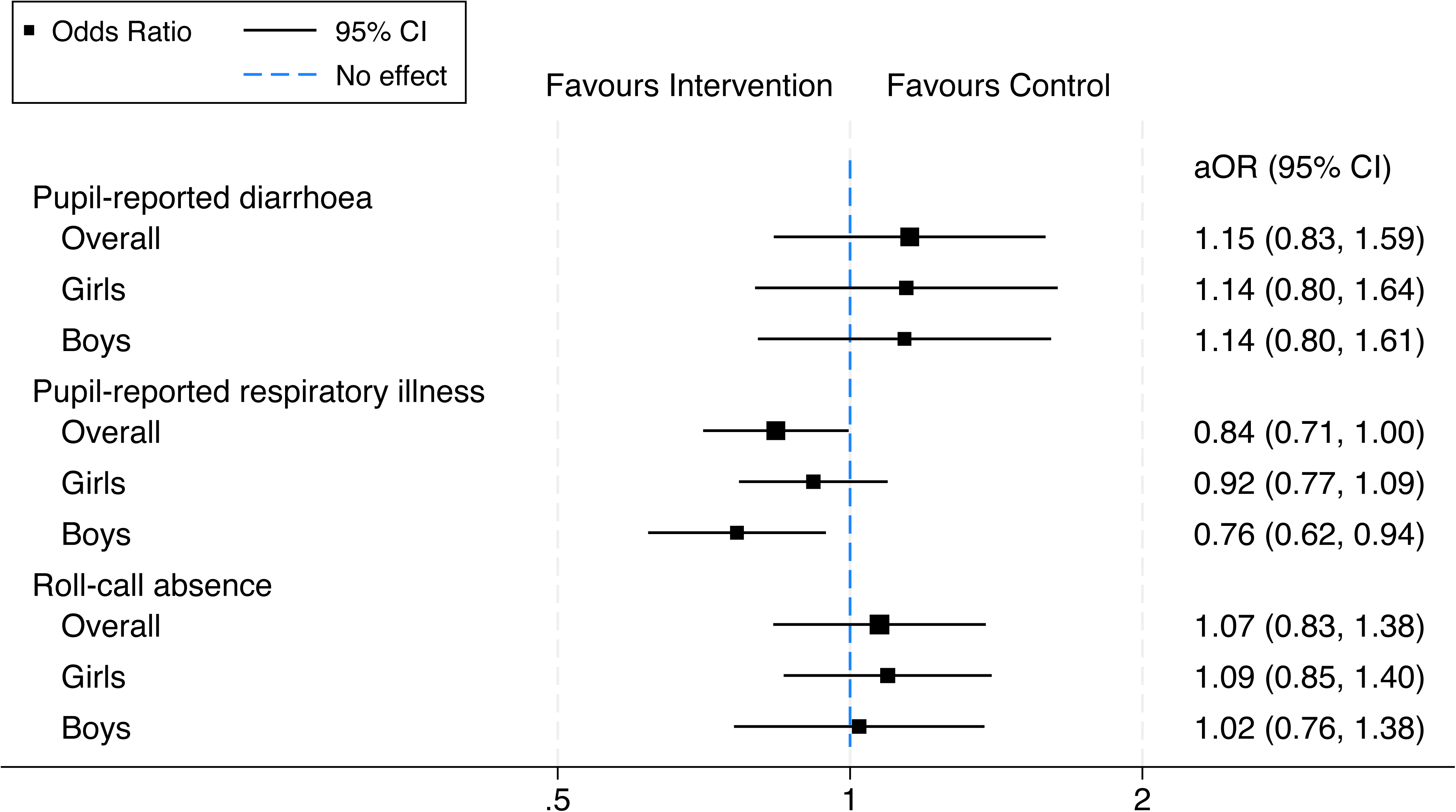
Gender-disaggregated intervention effects on primary outcomes.

Sensitivity analyses including all children in enrolled classes regardless of age, and adjusting for responsibility for household income generation (predictor of missingness at final follow-up) both produced similar findings (Supplementary Table 5).

## Discussion

In the WISE cluster-randomised trial, we found a borderline significant reduction in pupil-reported respiratory illness in the past seven days, and no evidence of reductions in diarrhoea or absence from school. These results point to the potential success of Project WISE at interrupting transmission of respiratory pathogens by increasing handwashing with soap, as hand hygiene interventions have been associated with reductions in risk of acute respiratory illness of 24% for school-aged children in LMICs.^2^ This finding is notable in the context of the COVID-19 pandemic, an active global threat during the study period (2021–2022), suggesting that school-based WASH can prevent disease amid social mixing at school, even when pupils have limited WASH access at home.

The lack of impact on pupil-reported diarrhoea may not be surprising given that schools received water and hygiene components but not intended upgraded toilet facilities within the study duration – access to clean sanitation facilities that safely remove excreta can be critical to interrupt transmission of faecal-oral pathogens.^30^ Providing safe, private spaces to change may be more important than menstrual products or sanitation technology in addressing menstruation-related absence^31^ – one explanation for the lack of effect on girls’ absence and the inconsistent effects on menstrual health outcomes, with only a small increase in menstrual care self-efficacy observed. School absence and wellbeing are multi-factorial; it seems feasible that the intervention (as received) may not have addressed enough factors to observe psychosocial and educational impacts.

The mixed effects of the WISE intervention on illness and absence are consistent with the existing literature,^9^ including multiple rigorous randomised trials.^19,32–34^ Impacts of WASH in schools are often context-specific and affected by factors such as local water access or underlying disease prevalence in the population.^9^ The high rates of past-week respiratory illness in this population (mean proportion of follow-ups with the outcome in the control arm 0·276) compared to other primary outcomes (0·073 and 0·103 for diarrhoea and absence, respectively) may have contributed to observing an effect for this outcome alone. We were unable to distinguish COVID-19 from other respiratory infections; it is unknown whether this high prevalence would persist in subsequent years.

Strengths of the study include the ‘open-cohort’ design that allowed minimal participant attrition, and absence triangulated through multiple measurement approaches – we provide evidence that pupil-reported absence is under-reported compared to roll-call absence. The use of pupil-reported measures for health outcomes is a limitation; using more ‘objective’ measures, such as stool-based pathogen detection versus self-reported diarrhoea,^35^ would help to minimise reporting bias, and enable differentiation of COVID-19 versus other respiratory infections. Concerns around bias in measuring illness among attendees are mitigated by the lack of differential absence rates between study arms, however it is possible some illnesses were missed by not following absentees. Follow-up was limited to one academic year for logistical reasons, so we are unable to evaluate the long-term impacts, or the additional effect of the sanitation component once received. With one borderline significant effect among three primary outcomes (without correction for multiple comparisons), we were unable to obtain strong evidence for the effectiveness of the intervention. For practical reasons we were limited in the number of schools that could be randomised; it is possible that the intervention had smaller effects only detectable with a larger cluster-randomised trial.

This study provides evidence that a school-based water and hygiene intervention implemented on a large scale across a large city can impact respiratory illness among schoolchildren, and demonstrated the utility of unannounced visits for absence tracking. However, the greater impact observed among boys remains unexplained, and future evaluations should include methods to differentiate pandemic and seasonal infection. Further research is warranted to establish the relationships between WASH-related illness and downstream educational outcomes – including illness-related and overall absence, educational progression, and gender parity in education – and strengthen understanding of the expected impacts of WASH in schools across multiple domains.

## Data Availability

All data produced in the present study will be made available in the London School of Hygiene & Tropical Medicine's Data Repository (https://datacompass.lshtm.ac.uk) with publication.

## Contributors

SB contributed to methodology, formal analysis, accessed and verified underlying data, writing – original draft, writing – review & editing, decision to submit. AE contributed to project administration, methodology, writing – review & editing. CO, WA, EH and EA contributed to methodology, writing – review & editing. BL contributed to methodology, accessed and verified underlying data, writing – review & editing. OC contributed to conceptualisation, methodology, writing – review & editing. RD contributed to conceptualisation, methodology, supervision, writing – review & editing, decision to submit. All authors approved the final version and accepted responsibility for publication.

## Data sharing

Deidentified participant data, a data dictionary defining each field in the set, and code to reproduce analyses using Stata will be made available in the London School of Hygiene & Tropical Medicine’s Data Repository (https://datacompass.lshtm.ac.uk) with publication.

## Acknowledgments

This study was funded by the Children’s Investment Fund Foundation, grant number 1907-03868. The authors would like to acknowledge the teachers, parents and pupils of the 60 schools in Addis Ababa who participated in the study and gave generously of their class time for this research. We also thank the *Splash* teams in Addis Ababa and Seattle for their assistance in the conduct of the study and coordination with schools. Lastly, we thank the survey staff from Holster International Research and Development Consultancy for their considerable efforts in data collection.

