## Supplementary Materials for "Impact of a school-based water and hygiene intervention on child health and school attendance in Addis Ababa, Ethiopia: a cluster-randomised controlled trial"

**Supplementary file A: Study protocol**

Protocol:

The WASH in Schools for Everyone (WISE) study: a randomised evaluation assessing the impact of a school-based water, sanitation and hygiene (WASH) intervention on child health and school attendance in Addis Ababa, Ethiopia.

S Bick, O Cumming, E Allen, and R Dreibelbis
London School of Hygiene and Tropical Medicine, UK

Version 9, 12/08/2021

**LSHTM Ethics Protocol 17761**

Lead PI:

Dr Robert Dreibelbis,
London School of Hygiene & Tropical Medicine
Keppel Street
London WC1E 7HT

London School of Hygiene & Tropical Medicine is the main research sponsor for this study. For further information regarding the sponsorship conditions, please contact the Research Governance and Integrity Office:

London School of Hygiene & Tropical Medicine
Keppel Street
London WC1E 7HT


### 1. Introduction

School-aged children in low- and middle-income countries (LMICs) are particularly susceptible to water, sanitation and hygiene (WASH)-related morbidities,[1] including gastrointestinal[2, 3] and respiratory infections,[4] and soil-transmitted helminth (STH) infection,[5] due to frequent social mixing and poor hand hygiene.[6, 7] These health risks are associated with absence from school,[8] which can in turn act as a measure of health status.[9] The effects of morbidity and absence include lower test scores[10, 11] and dropout,[10, 12] particularly in low-resource settings, with implications on social, occupational and health outcomes associated with continued education.[13-15] While reasons for absence and dropout are varied,[12] inadequate WASH conditions in schools may present barriers to attendance – through lack of services, especially for menstruating girls, and increased illness,[16] with latrine cleanliness a significant factor.[17] Pupils' academic performance also may be affected by dehydration where there is inadequate water supply.[18]

Girls in LMICs are disproportionally affected by poor access to sanitation,[19] potentially accounting for gender disparities in enrolment and attendance; despite significant progress globally, girls' participation rates remain lower than those of boys in 53 LMICs.[20] Pubescent-age girls face greater challenges in accessing sanitation, including threats to privacy and safety.[21, 22] Menstruation cyclically increases the need to access sanitation,[23] and low quality and availability of water, limited health and sexuality information, and lack of hygienic materials, disposal facilities and privacy leave girls with limited options for menstrual hygiene management (MHM),[19, 24] resulting in psychosocial stress,[25-27] increased risk of urogenital infection[28] among other health risks, and impacting school participation and attainment.[29-31] Girls may fall behind and drop out,[32] and girls' enrolment is negatively associated with onset of menarche[33] and has been seen to decline post-menarche where there is limited water access.[34] Improving WASH in schools may therefore also contribute towards attaining gender parity in educational outcomes.

Although WASH in schools (WinS) interventions have been hypothesised to improve children's health outcomes, evidence of their impact has been mixed. Handwashing interventions have been shown to reduce the incidence of diarrhoea by 53%–73%.[35] A recent systematic review of WinS intervention studies in low-income countries[36] found significant reductions in pupil-reported diarrhoeal disease between 29% and 50%,[1, 37] reduced incidence of respiratory illness,[1, 38, 39] and reduced STH incidence[40] and reinfection,[41] following varied WinS interventions including provision of water for drinking and handwashing, water quality, sanitation, and hygiene promotion. Other studies found no significant impacts on diarrhoea[39, 42] (although low diarrhoea rates in the population prevented detection of differences), STH prevalence[43] or respiratory illness,[42] or saw positive impacts for only some of their disease outcomes.[39, 41, 44-48] Outcomes were affected by contextual factors, for example, two studies noted an effect of local water availability on reduction in diarrhoeal disease.[3, 47]

Evidence of the impact of WinS interventions on educational outcomes has been similarly ambiguous.[36, 49] WASH improvements have been shown to reduced absenteeism,[38, 42, 50-52] but not universally: some studies observed no differential attendance between intervention arms,[1, 53-55] but specific effects on girls' absence alone,[55] or on absence due to diarrhoea.[1] Only one[42] of five randomised controlled trials (RCTs)[42, 53-56] reports significantly lower overall absence rates. Drinking water provision in schools improved students' hydration status,[18] and has resulted in attendance improvements in dry seasons.[52] WinS interventions have been shown to increase enrolment and enrolment parity,[57] and girls' attendance was seen to benefit specifically from same-sex toilets in schools.[21] Evidence from MHM interventions is more limited: school-based sanitary pad provision and hygiene education reduced absenteeism in a pilot study,[58] but no effects on attendance or dropout were seen following other sanitary product programmes.[59, 60] Perceived lack of privacy, not toilet type or cleanliness, has been linked to girls' absence, associated with a 2.6 greater odds of absence during menstruation;[61] providing safe, private spaces to change may be more important than products or sanitation technology in addressing menstruation-related absence.[62]

In 2012, UNICEF estimated that 49% of schools in low-income settings had inadequate access to water and 55% had inadequate access to sanitation.[63] Access to WASH facilities in school environments is included in the Sustainable Development Goals (SDGs)[64] and is essential in ensuring dignity and equity, and promoting women's equality and empowerment. To achieve these goals, interventions must be accompanied by sustained management of services over time.[43, 65, 66] While construction of latrines, and other improvements to infrastructure, can increase their use,[67] increased exposure to faecal pathogens may result if they are poorly maintained, or lack consistent availability of soap and water for practicing handwashing.[3] Several publications highlight that combined interventions of multiple components are more effective than single interventions at reducing disease and absenteeism.[3, 54, 55] WinS evaluations have noted challenges of intervention fidelity and adherence,[68-70] and demonstrated the impact of adherence on health outcomes.[70] Improved understanding of the health and educational outcomes of school-based WASH interventions, and factors that contribute to their success, can help prioritise resources and design better interventions to improve children's futures.

### 2. Study rationale

The WASH in Schools for Everyone (WISE) programme provides an opportunity to strengthen the body of knowledge around the impact of WinS on child health, school attendance and gender parity in educational outcomes. Systematic reviews of the impacts of WASH in LMIC schools on health and educational outcomes,[36] school-based hand hygiene on disease outcomes,[35] and MHM interventions[71] and school sanitation[49] on absence and psychosocial outcomes all point to the paucity of rigorous evidence for the effects of WASH in schools. Absence intervention studies in particular have suffered from high risk of bias,[49] necessitating more robust studies, especially large-scale randomised controlled trials. While evidence for impacts on diarrhoea is more established, broader educational outcomes are harder to assess. Considering the validity of unannounced attendance spot-checks over school records alone (as utilised previously),[1, 23, 54, 72] we will triangulate data from multiple sources, including detailed absence measures in unannounced visits for students. Few studies link absence to WASH-related disease; this study will assess illness on the pathway to absence, and include further assessment of education participation and psychosocial outcomes.

The few intervention studies that include a focus on MHM in schools in LMICs mainly use qualitative, participatory and/or descriptive methods.[19] Quantitative studies of the impacts of MHM on education are scarce, and have mainly considered absorbent type[31, 58, 61]. Moreover, they often assess one aspect of the full MHM definition, which includes frequency of absorbent change, washing of the body, adequate disposal and privacy.[73] Negative psychosocial outcomes of poor MHM have also been well documented in qualitative studies.[74] The proposed evaluation offers the opportunity to quantify the impacts of MHM on absence and psychosocial outcomes including menstrual hygiene self-efficacy, in the context of a WASH intervention in schools in LMICs. Sub-population analysis of evaluation outcomes in girls post-menarche will allow assessment of the gendered effects of WASH improvements.

The WISE evaluation described here will evaluate the effects of a comprehensive WASH intervention, including water and sanitation infrastructure, behaviour change promotion and targeted MHM services, on child health and attendance in schools in Addis Ababa, Ethiopia.

### 3. Study setting

Addis Ababa is the capital and largest city of Ethiopia, and as such has greater coverage of water and sanitation in schools compared to rural areas[75]. 46% of schools in Ethiopia have a year-round onsite water source[76], but only 6% had access to basic hygiene services in 2016[77]. In a survey of Ethiopian schoolchildren, 95% of girls said they did not have access to safe toilet facilities with water access at school[78]. 97% of boys and 89% percent of girls have ever attended school in Addis Ababa, for an average of 8.3 years for boys and 6.9 years for girls[79]. Although the gender gap in primary school enrolment in Ethiopia has narrowed, from around 15% in 2000 to 4% in 2011, net enrolment at the secondary level has not significantly altered[80]. Factors outside education including perceptions about earning potential, and favouritism of boys contribute to pervasive gender disparities in education[81]. The average age of menarche of Addis Ababa schoolchildren is an estimated 13.7 years[82].

The US-based NGO *Splash* will deliver comprehensive school-based water, sanitation, and hygiene improvements to all schools in Addis Ababa, Ethiopia over a 5-year period. Eligible schools are public schools in Addis Ababa, including kindergarten, primary, and secondary schools and excluding rented or temporary schools serving less than 50 students. There are around 460 schools serving over 420,000 children. This project aims to reach 100% of these schools through delivering the intervention to pre-defined groups of schools on an annual basis.

#### Disruptions due to the COVID-19 pandemic

The study was originally planned to begin in March 2020, but the COVID-19 pandemic led to school closures in Ethiopia and limited travel. As of January 2021, schools have re-opened in Addis Ababa. The following study procedures have been adapted to the COVID-19 situation, and include measures to minimise risk to the study of further school closures, and include respiratory illness among students as a primary outcome.

### 4. Research aims and objectives

The aim of this study is to assess the effects of school-based, comprehensive water, sanitation, and hygiene improvements on child health and educational outcomes in schools in Addis Ababa, Ethiopia.

Specific objectives are to evaluate:

1. The effect of the intervention on pupil-reported diarrhoea and respiratory illness in the last 7 days, and other relevant health outcomes (described below).
2. The effect of the intervention on pupil attendance and causes of absence.
3. The effective of the intervention on pupils' subjective wellbeing.
4. The effect of the intervention on educational and MHM outcomes among girls, by assessing:
    a. Gender parity in enrolment and attendance.
    b. Absence during menstruation among girls aged 10 and above.
    c. Education participation and subjective wellbeing among girls aged 10 and above.
    d. MHM self-efficacy and perceptions of met needs among girls aged 10 and above.
5. The cost-effectiveness of the intervention.
6. Intervention fidelity using process evaluation methods.

### 5. Methods

#### Evaluation overview

The proposed study is a cluster-randomised controlled trial (cRCT) design. The prior version of the approved protocol utilised a non-randomised stepped wedge evaluation. However, due to changes in the implementation timeline due to COVID-19 and uncertainty around future implementation groups, the opportunity to transition the study to a cluster-randomised trial was identified.

In order to account for further disruption to implementation schedules and loss to follow-up, schools will be over-sampled to ensure that adequate power is retained for analysis of the primary outcomes. 30 schools will be randomly allocated to intervention and control arms and enrolled in the study, ensuring retention of minimum 25 schools in each arm if 17% of schools are lost to follow-up.

#### The WISE intervention

The WISE intervention will be delivered in annual implementation groups, consisting of 100 to 200 schools each academic year (Sept – May). Each annual implementation round (referred to as "Groups") is further divided into three batches.

Approximate dates are listed below:

| Batch 1 | Batch 2 | Batch 3 |
|---|---|---|
| Sept – Nov | Dec – Feb | Mar – May |

*Splash* is currently preparing the list of schools that will be included in the 2021/2022 and 2022/2023 academic years implementation schedules. LSHTM will collaborate with *Splash* to randomly allocate 30 of these schools to be part of the 2021/2022, batch 1 implementation and 30 schools to be part of the 2022/2023 implementation (any batch).
Schools will constitute the study clusters for data collection. Since class sizes were approximately 20-30 students in the pilot study, four sentinel classrooms of students (approx.100 students in total / school) will be randomly selected as study participants at each of these schools at the start of data collection.

A stratified sampling approach will be used during allocation of schools to intervention and control arms to ensure balance in school size and schools with a kindergarten between trial arms. If there are fewer than 20 schools with a kindergarten in each trial arm necessary for the kindergarten sub-study, additional kindergarten schools will be sampled to meet this requirement. We estimate that approximately 40 schools will be enrolled in each trial arm: 30 in the older children study, and an additional 10 kindergarten schools not part of the older children study.

The Consort Diagram is attached below:

**WISE Study Design Flow Diagram**

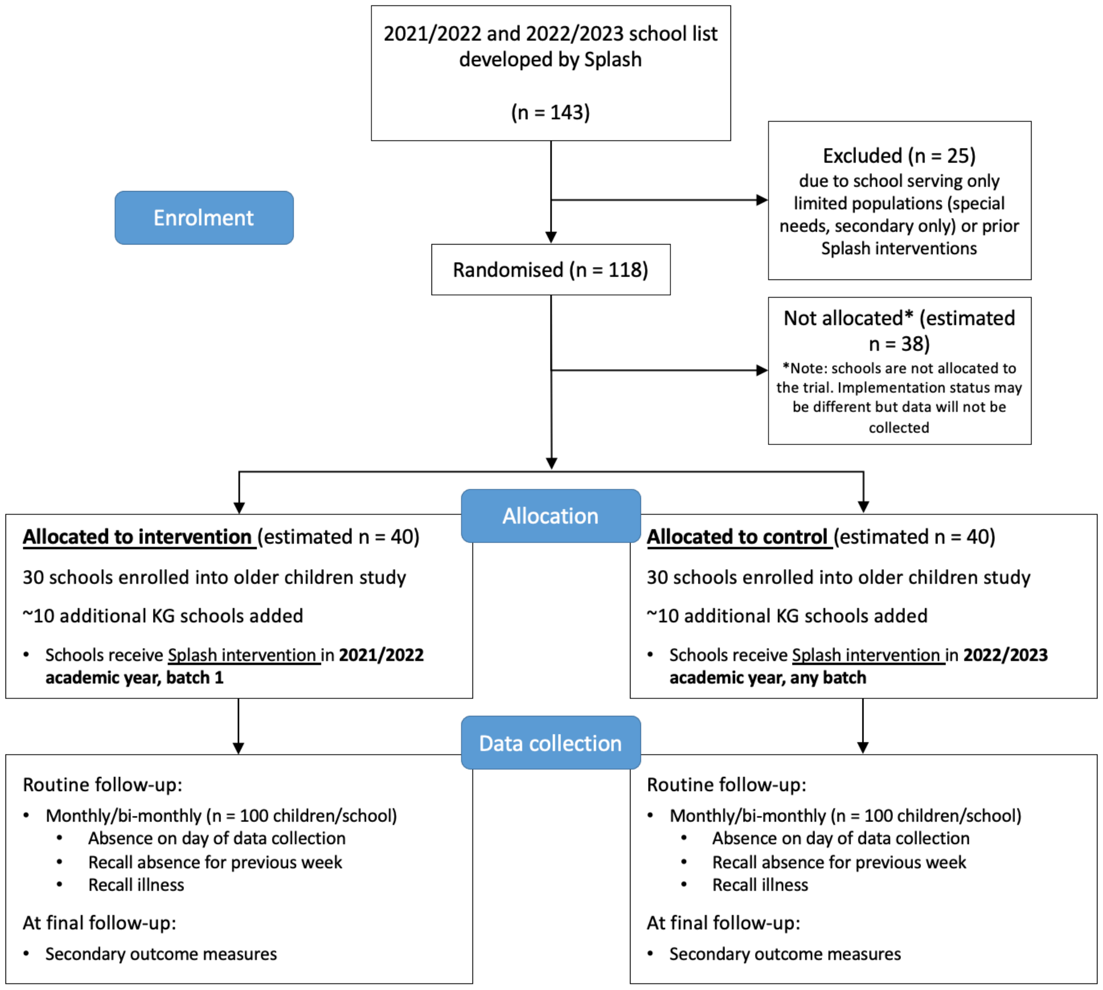

### Evaluation outcomes

#### *Primary outcomes*

The primary outcomes are:

1. Prevalence of self-reported diarrhoeal disease and respiratory illness in the past 2 and 7 days; and
2. Absence calculated as i) proportion of students absent during routine follow-up visits, and ii) cumulative absence per child per school year.

#### *Secondary outcomes*

Secondary outcomes that will be evaluated include:

1. Subjective well-being assessed using a smiley faces visual analogue scale, Strengths and Difficulties Questionnaire,[83] and a measure of sanitation-related quality of life (Ross; publication pending).
2. MHM self-efficacy and menstrual perceptions, based on scales developed in Bangladesh (Hunter; publication pending) and Uganda;[84]
3. Gender parity in school enrolment, using the adjusted gender parity index,[85] based on school rosters; and
4. Cost-effectiveness, measured by incremental cost per case / absence day averted by the intervention.[86]

### Data collection approach

The following data collection activities will be conducted in the four selected sentinel classrooms in each school:

1. A detailed **enrolment survey** will be conducted at the start of the academic year (Sept):
   - Demographic information and household WASH access
   - School WASH conditions
   - **Primary outcomes of self-reported absence and illness (diarrhoea and respiratory illness)**

2. These same students will then be followed up over the academic year with unannounced **monthly visits** approximately every 4–6 weeks:
   - Attendance on day of data collection
   - Self-reported absence and illness in past week (diarrhoea and respiratory illness)
3. At the final monthly follow-up, additional data will collect data on secondary outcomes:
   - Subjective wellbeing and sanitation-related quality of life (all students), the strengths and difficulties questionnaire (over age 11 only), menstrual perceptions and MHM self-efficacy (girls over age 10 only)

### Rationale

Assessing the primary outcome of absence using longitudinal data with unannounced visits provides a more complete assessment of absence rates over the course of the year, and avoids issues around assessing attendance among students already in attendance. Cumulative attendance is likely a more important predictor of educational attainment than absence at one point in time. As the secondary outcomes are continuous measures, the study is well powered to detect small changes in secondary outcomes using data from annual time points alone (see sample size calculation section). The final follow-up survey will be shorter, require in-depth surveys only with the relevant groups of older students, and will therefore be less disruptive to the whole school during exam season.

### Interventions

The WASH in Schools for everyone (WISE) programme, implemented by *Splash*, aims to improve child health and attendance by 1) improving WASH infrastructure in schools, 2) promoting WASH behaviour change and 3) strengthening MHM services targeting girls aged 10 and above, across government schools in Addis Ababa. The intervention includes:

WASH infrastructure:

- Water storage tanks and water filtration systems
- Durable plastic drinking water and handwashing stations (Figure 1)
- New/rehabilitated toilet facilities, that are gender-segregated, separate for students and teachers, wheelchair-accessible, well-lit and ventilated, able to close and lock from the inside

Behaviour change promotion:

- Training of focal teachers to champion WASH programme and organise 20–30-student ‘hygiene club’
- Annual soap drive and restocking of soap
- Behavioural ‘nudges’, including posters and mirrors at handwashing stations, and hygiene club students with colourful vests to stand near handwashing stations

MHM services:

- Toilets have strong, durable door with lock, adequate lighting in stall and hallway, water tap and bucket and waste bin
- Training of focal teachers to organise student ‘gender clubs’
- Emergency menstrual hygiene products available at school
- Education on puberty and menstruation for all pupils above 10
- Q&A session for girls above 10 and demonstration of products
- Peer mentoring of younger girls by older girls
- Distribution of parent resource guide on MHM
- School-wide celebration of menstrual hygiene day

*Figure 1: Splash* drinking water and handwashing stations.

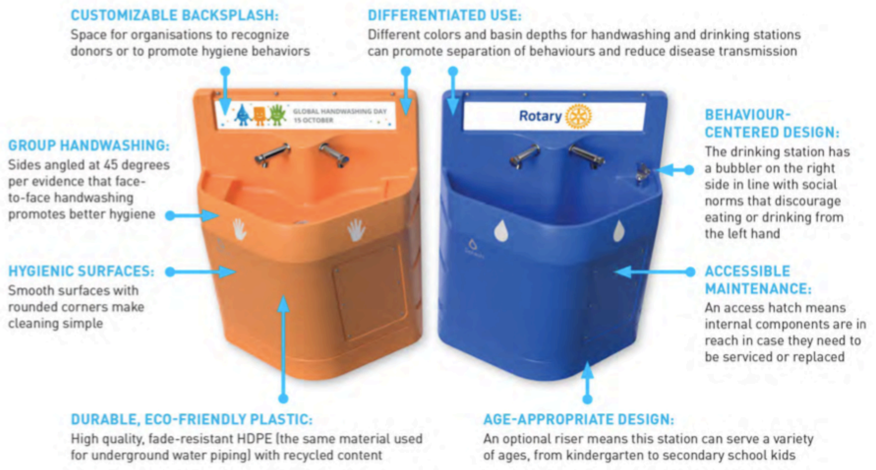

### Inclusion and Exclusion Criteria

#### *School criteria*

Inclusion:

- School is part of appropriate *Splash* Project WISE implementation groups
- School agrees to participate in the research

Exclusion:

- School has received a water, sanitation, or hygiene intervention in the three years prior to study activities
- School is kindergarten only
- School is secondary only (standards 9 and above, equivalent)
- School provides education to vulnerable population groups only

#### *Child criteria*

Inclusion

- Child is registered at a school that meets eligibility criteria and selected for the study
- Child is in grades 2-8
- Child's parents do not return the opt-out consent form (see below)
- Child provides assent to participate in the research

Exclusion

- Child is unable to provide assent for data collection
- Child is 17 or older (excluded from analysis)
- Child does not meet age eligibility for specific data collection activities (see below)

In the pilot study (see next section), there was considerable variability in the ages of students in grades 9-10, with approximately half aged 17 or older. Inclusion was therefore restricted to schools with a primary school (grades 1-8). Students 17 or older in the selected classes (estimated 6% of students) will be included in data collection in order to minimise social exclusion, but excluded at the analysis stage.

### 6. Study Procedures

#### Study pilot

Data collection tools and methods for school and student enrolment, follow-up surveys and staff surveys were piloted in 5 randomly selected schools not included in the primary impact evaluation but where *Splash* implemented the WISE intervention in 2020/2021, between May and July 2021. During piloting, in-country data collection partners, in collaboration with LSHTM researchers, followed full study procedures outlined below, with one minor variation: follow-up of selected sentinel classrooms occurred roughly 3 weeks after enrolment and only one round of follow-up occurred.

*Piloting study procedures:*

Prior to data collection, a member of the data collection team approached each school, explained study procedures to the school principal, and held parent meetings to explain the purposes of the study and distribute parental opt-out forms. This data collector also collected a copy of the school roster to obtain enrolment data.

Approximately 1 week later, data collection teams returned to the school to test student enrolment data collection procedures. Data collection staff obtained oral assent from all students and verified eligibility. Data collection staff implemented enrolment surveys with students in two selected classrooms. Data were be collected on mobile devices by survey teams in the field, and were be uploaded to servers each evening, or whenever internet connection allowed. A member of the data collection team returned to the school approximately 3 weeks later to test the follow-up survey (not including secondary outcome measures). Further details on each of these procedures are described below.

The study pilot will be used to assess the overall logistics of field data collection and develop appropriate systems for data management and coordination. The study pilot will also provide an opportunity to verify assumptions included in sample size calculations.

*Tool adaptation*

We will test and adapt MHM self-efficacy and menstrual perception scales. Tool adaptation will follow standard approaches to revising and adapting data collection instruments and will occur after the piloting period but before the final follow-up when these measures are required. For tool adaptation, we will complete a series of focus group discussion (FGDs) with girls attending pilot schools during the 2021/2022 academic year. During FGDs, researchers will present children with the questions included in the MHM self-efficacy and menstrual perception scales and discuss participant understandings of the questions and pre-defined response categories. Tools will be adapted to ensure that questions are understood by participants and that items have been translated appropriately.

Questions will also be adapted to reflect local MHM practices and preferences – such as the brand and/or generic names for specific absorbent materials and local terminology related to menstrual hygiene. Directions to respondents – including both paper-based and orally-administered instructions – will also be discussed with participants. Data collection tools will be adapted based on participant feedback. We anticipate that we will complete approximately 4 FGDs with a maximum of 8 participants each, for a total enrolment of 32 students. We will purposively sample students to ensure that FGDs capture the range of student ages (early adolescent, late adolescent, secondary schools, etc.). Consent and assent will follow the same procedures outlined in the rest of the study.

### Main study

#### *Enrolment survey*

The enrolment survey will be completed at the start of follow up in each participating school with students from the four sentinel classrooms (12pprox.. 100 students in total). The enrolment survey will include the following:

- All students:
    - recall of full- and partial-day absence over the preceding 5 school days, alongside information recorded about causes of absence
    - recent illness through self-reported health outcomes over the preceding 2 and 7 days, including stool frequency and other "highly credible" symptoms of enteric infection,[87, 88] and symptoms of other respiratory disease outcomes (coughing, sneezing, etc.)
    - Basic information on household socio-demographics, WASH conditions in the home, and student involvement in key activities known to impact school attendance, such as domestic responsibilities (childcare, chores) and income generation activities.

Enrolment surveys will be completed one-to-one on tablets.
During enrolment, one enumerator will collect a copy of school rosters to assess yearly enrolment across the school. This will be updated with school staff at the end of the academic year.

#### *Follow-up surveys*

Routine absence tracking for enrolled students will take place every 4-6 weeks. Data collection schedules will be managed in direct partnership between LSHTM and in-country partners to ensure sufficient variability between follow up days and times. Data collection staff will visit sentinel classrooms and record attendance on the day of data collection using written class rosters. At each round of data collection, data collection staff will complete a brief orally-administered survey (< 5 min) using mobile devices with each student to assess recent absence and causes of absence. The first follow-up visit will also be an opportunity to conduct enrolment surveys with any students who were absent on the day of the enrolment survey.

*Final monthly follow-up*

The final monthly follow-up of enrolled students enumerators will follow all procedures described above as part of routine follow-up surveys. In addition, the following data will be collected:

- All students:
  - subjective well-being, measured by children using a visual analogue scale (smiley faces)
- Students age 11 and older:
  - Strengths and Difficulties Questionnaire (SDQ-25), a widely used measure of student behavioural and mental health challenges that is designed for use among school-aged children and has been used in a number of Sub-Saharan African countries
  - Sanitation Quality of Life measure (SanQoL) based on newly developed measures based on adult populations in Mozambique. This five-item measure will be tested and adapted during the study pilot
- Girls aged 10 and older:
  - Self-administered surveys that include questions on MHM-related psychosocial outcomes, including measurements of menstrual hygiene self-efficacy, menstrual practices and menstrual perceptions (Menstrual Practice Needs Scale) – a measure of unmet menstrual needs from the perspective of the girls themselves. Both tools have previously been used among school-going populations. These measures, combined with subjective well-being and measures of education participation, will provide a more comprehensive assessment of the impact of the WISE intervention on girls than focusing on attendance-related outcomes alone. Surveys include visual analogues, and will be calibrated using simple questions about level of confidence.

Sections of the survey for older students will be self-administered on paper forms, with instructions given to small groups of students. Data from these written components will then be entered into tablets following data collection.

Data collection in enrolled schools is anticipated to start at the beginning of the 2021/2022 academic year, assuming all necessary approvals are in place, study tools have been piloted, and parents have been notified of the study. The anticipated start date is September 2021.

Primary impact of the intervention will be assessed based on data collected after the *Splash* intervention has been completed in intervention schools. Because the exact implementation dates are determined by *Splash* and the participating schools, some monthly data collection in intervention schools may occur before and/or during active implementation of the full *Splash* intervention. Any data collected prior to and/or during the active *Splash* intervention will not be included in the primary impact evaluation analysis. We anticipate that for intervention schools, this will allow 4 – 6 months of active follow-up after implementation is completed.

A sample timeline for data collection is presented below:

| | 2021 | | | | | | | | | | 2022 | | | | |
|---|---|---|---|---|---|---|---|---|---|---|---|---|---|---|---|
| | J | F | M | A | M | | S | O | N | D | J | F | M | A | M |
| **Data collection** | | | | | | | | | | | | | | | |
| Ethics | ■ | | | | | | | | | | | | | | |
| Pilot testing, sample size revision | | ■ | ■ | ■ | | | | | | | | | | | |
| Enrolment survey | | | | | | | ■ | | | | | | | | |
| Follow-up (every 6 weeks) | | | | | | | | ■ | ■ | ■ | ■ | ■ | ■ | ■ | |
| Final follow-up (Secondary outcomes measured) | | | | | | | | | | | | | | | ■ |
| **Engagement** | | | | | | | | | | | | | | | |
| Joint planning with BOE, SPLASH and CIFF | | | ■ | ■ | ■ | | | | | | | | | | |
| **Implementation** | | | | | | | | | | | | | | | |
| Group 3, Batch 1 | | | | | | | ■ | ■ | ■ | | | | | | |

*Routine data collection*

Routine data collection and monitoring systems implemented by *Splash* will include an assessment of cleanliness and maintenance of WASH infrastructure, as well as data on costs and quantities of resources used. These outcome measures will be used to conduct a process evaluation and will be used in calculations of cost-effectiveness.

Routine data collection will also include periodic surveys / semi-structured interviews with the school principal and one or more teachers/school staff per school on a rotational basis over the course of the evaluation. Data collected from these surveys will include previous WASH intervention, engagement with the implementing partners, direct involvement in intervention activities, and costs / expenditures. A brief facility inspection will also be included periodically at follow-up up visits.

### 7. Sample size calculation and analysis

#### Analysis

All data will be analysed using Stata[89] and/or R[90]. Primary impact measures of absence and diarrhoea will be as cumulative number of days absent / 100 school days after adjustment for child- and school-specific covariates. Secondary outcomes of well-being, education participation, cause of absence and gender ratios in enrolment will be compared between trial arms. Our primary assessment of cost-effectiveness will utilise standard costing and cost-effectiveness measures, e.g. incremental cost per case / absence day averted by the intervention.[86] Costing will follow the reference case of the global health cost consortium,[91] with cost data collected during routine data collection.

#### Power and sample size calculations

Power and sample size calculations were conducted to assess changes in absence in follow-up visits to two sentinel classrooms in each school (approximately 100 students), and were based on 25 clusters in each study arm to account for potential loss to follow-up or disruption to implementation schedules. We assumed a conservative mean absence rate of 11% throughout the follow-up period in the control schools based on available longitudinal data. Estimates were taken from unpublished partner data taken from a longitudinal cohort of children attending mixed education primary and secondary schools in peri-urban Indonesia and likely an underestimation of the true absence rate in this population.[54, 92]

Minimum detectable difference between control and intervention schools on the average absence rate or average illness longitudinal prevalence over the follow-up period is determined by the standard deviation of mean absence rate or mean longitudinal prevalence of illness and the intra-cluster correlation coefficient (ICC), both of which are unknown.

Assuming mean absence rate 11% (SD 7%), ICC 0.15, and minimum 25 clusters in each study arm, absence data from approximately 100 children per school is sufficient to detect a 20% relative reduction in the mean absence rate.

For student illness, assuming a longitudinal prevalence of illness of 8% over the cumulative follow up period (SD 5%), ICC 0.15, and minimum 25 clusters in each study arm, absence data from approximately 100 children per school is sufficient to detect a 20% relative reduction in the mean longitudinal prevalence of illness.

In the pilot study, school-level ICC values for measures of pupil illness were estimated to lie in the range of 0.07 to 0.1. The sample sizes calculated are therefore suitable conservative estimates.

Secondary outcome measures are limited primarily to girls who are aged 11 or older. We made the assumption that only 1 out of every 3 schools included in the evaluation will include a randomly selected classroom with girls aged 11 or older, that only 1 classroom with girls aged 11 or older will be selected in these schools, and that girls will make up 50% of the class for a total sample of 25 girls from 8 schools in the intervention and 8 schools in the control arms. We assumed a mean score of 1.76 (SD: 0.34) on the menstrual needs scale in the control group based on unpublished data from similar populations. Assuming an ICC of 0.15, our study is designed to detect a 12% difference in mean scores between control and intervention arms. For the menstrual hygiene self-efficacy score, we assumed a mean score of 69.6 (SD: 16.6) based on unpublished data from both tools in similar urban, resource limited environments. Assuming an ICC of 0.15 our study is designed to detect a 14% difference in mean scores between control and intervention arms of the study.

### 8. Special considerations for Kindergarten (KG) students

Due to the high burden of diarrhoeal disease concentrated in young children but considering that young children cannot self-report diarrhoea outcomes, additional study procedures have been added to better document impact among young children. This will follow the same cRCT design as the main study but include a subset of study clusters, those included in the older children study that have a kindergarten. A stratified sampling approach will ensure that a sufficient number of schools (20 in each trial arm) with a kindergarten are allocated. Outcomes will be longitudinal prevalence of diarrhoea and attendance, assessed through telephone interviews with parents of 20 randomly selected kindergarten students.

### Study pilot

All data collection tools and methods will be piloted in 3 randomly selected schools with a kindergarten not included in the primary impact evaluation but where *Splash* implemented the WISE intervention in 2020/2021, and will take place over an approximately one-month period. During piloting, in-country data collection partners, in collaboration with LSHTM researchers, will follow full study procedures outlined below, with one minor variation: instead of 4 weekly telephone calls to parents, only 2 total phone calls will be made.

*Piloting study procedures:*

Prior to data collection, a member of the data collection team will collect contact information for parents of the 20 randomly selected students from the school. Data collectors will call parents once a week for 2 consecutive weeks. Each call, data collectors will collect consent from parents, and conduct a short survey about the number of days in the past week their child has been ill or absent from kindergarten. During the first call, they will also collect demographic and household WASH information.

The study pilot will be used to develop appropriate systems for data management and coordination. The study pilot will also provide an opportunity to verify assumptions included in sample size calculations.

### Main study – Telephone interviews

Approximately 6 months into the academic year (~ Feb/March 2022), data collectors will obtain the contact details for parents of 20 randomly selected students from the 40 schools. Data collectors will call parents once a week for 4 consecutive weeks. Each call, data collectors will collect consent from parents, and conduct a short survey about the number of days in the past week their child has been ill or absent from kindergarten. During the first call, they will also collect demographic and household WASH information.

*Enrolment and Consent Procedures*

Verbal consent will be collected from parents contacted via telephone (script attached), and enumerators will document the consent processes and provide a written copy of the agreed consent document to parents following data collection.

### Sample size calculation and analysis

Power and sample size calculations were conducted to assess changes in the longitudinal prevalence of reported diarrhoea from telephone interviews over one month. For sample size calculations, we assumed a mean longitudinal prevalence of 8% (SD 5%) among pupils in the control group and an

ICC of 0.15. For 20 clusters from each study arm, diarrhoeal longitudinal prevalence data from approximately 20 children per school is sufficient to detect a 24% reduction in diarrhoea.

### 9. Participant enrolment and safety

#### Participant enrolment

Local staff will engage selected schools, holding meetings with school leadership and providing a full explanation of the intervention and study. School principals will receive a full orientation including explanation of research questions, consent processes, anonymisation and guidance for communicating to parents, in order to improve participation in the study and ensure parents and students understand the research.

Focal teachers and maintenance staff will be identified for training. Consent / assent will be obtained in multiple ways:

1. Written consent will be obtained from the school principal/headteacher at each participating school to cover all data collection. The principal will sign a *loco parentis* document, verifying that he/she understands the proposed study and granting permission for data collection to commence at the school.
2. COVID-19 restrictions permitting, the purposes of the study and study procedures will be explained at a parent meeting at the school prior to data collection. Parental opt-out forms will be distributed to all students at the school, alongside information on the nature of the research and explaining that there are no risks associated with participation or penalties for non-participation. Parents will be given the opportunity to return forms to the school before data collection. If attendance at the parent meeting is below a certain threshold determined in partnership with local collaborators and in country ethical reviews, we will hold multiple parent events to ensure all parents are informed and have the chance to ask questions before making a decision. For parents contacted by telephone as part of the kindergarten impact assessment, verbal consent will be collected prior to data collection. If schools are unable to hold in-person meetings due to COVID-19 restrictions on adult gatherings, we will instead send two or more letters home with students to provide parents with increased opportunities to learn about the study. We will also utilise any specific systems the local school has to communicate with parents – such as newsletters, bulletin boards, or other communication means. All communication materials will include the number for the local data collection partner who can answer any questions parents have about the study.
3. Oral assent from participating children will be recorded prior to each data collection activity. Forms written in very plain language will be given explaining the nature of the data collection activity, and pupils assured that their participation is voluntary and without risk.

Study participants may withdraw from the study at any point. Withdrawal from the study after enrolment will lead to no further analysis, but data already collected and analysed will be used, unless requested otherwise by participants. No incentives will be given to study participants.

### Participant safety

School children may feel uncomfortable discussing topics such as menstruation or sharing information on cause of absence for fear of repercussions. Participants will be assured of the confidentiality of any information they provide and that they are able to suspend participation at any time with no consequence to themselves. Surveys on MHM-related outcomes will be self-administered paper surveys that girls can complete these questions without fear of the enumerators knowing their responses.

School teachers and principals may feel uncomfortable having their facilities monitored if they are in poor condition, and may be concerned about this information being distributed. All procedures will be explained in depth and steps will be taken to address any concerns felt by participants.

In the event that any child becomes ill or upset during any data collection activity, the activity will immediately stop, and field staff will take necessary action to ensure the participant received appropriate counselling or medical support. The methodology would be reviewed to determine how it may be revised to avoid harm. If a child discloses that they or someone they know is at risk during the interaction with researchers, the enumerator will take necessary action to report the child’s situation to the relevant authority. The enumerator will inform the child of their obligation to do so and will ensure that the child receives appropriate counselling and/or medical support. Researchers will receive training prior to commencing the study to enable them to best approach such situations.

COVID-19 safeguards are in active development with both implementing agencies and data collection partners and will be done in-line with local guidelines for data collection.

COVID-19 prevention measures will include temperature checks and provision of face masks and hand sanitiser to enumerators before entering a school. Enumerators will maintain physical distancing at all times and will conduct interviews in well-ventilated/outdoor areas. Data collection will be conducted in accordance with all national COVID-19 guidelines and any additional requirements by data collection partners. A complete COVID-19 risk management strategy will be shared prior to the start of any data collection activities.

### 10. Study limitations

There are a number of limitations to the study design. Firstly, that it is impossible to blind any participants to their allocation in the intervention or control arms at any point in the study, leading to possible overestimated effects of the intervention. Additionally, the menstrual hygiene education component may lead to reduction of some of the stigma attached to menstruation in intervention schools, leading to differential responses as girls may share more freely about MHM practices and perceptions, or menstruation-related absence. In order to mitigate this, field staff will emphasise to all students the purposes of the study to inform rather than judge or inspect their attendance record. The use of self-reported outcome measures may be prone to recall and social desirability biases, and varied interpretations of the definition of “diarrhoea”[68]. The pupil survey will include a clear definition of diarrhoea in terms of stool frequency to improve the validity of this measure.

The main envisaged risk to the study is low participation of schools in the control group in the study, due to no perceived benefit of participation over a year prior to receiving the intervention. Delays to implementation may also result in schools being moved between intervention and control arms. The flexible nature of the study design with repeated measures will allow the precise time schools switch to intervention to be captured and adjusted for in analysis. There is also the possibility of low intervention fidelity, adherence and maintenance over the course of the study period. *Splash* will work closely with participating schools to monitor the implementation of the intervention, and the process evaluation included in this study will assess the extent to which the intervention has proceeded as planned.

### 11. Confidentiality

Personal identifiers collected with the survey, including names and telephone numbers, will be stored separately from other, de-identified data and removed from electronic data records and analysis. Physical forms containing personal identifiers will be kept in a locked office and safely disposed of after the study is complete. Electronic data will be encrypted and stored on secured, password-protected electronic databases to prevent release of information.
Data may be made available in the public domain after the study has been completed. Various safeguards will be enacted to ensure that no school or individual child is directly or indirectly identifiable. Explicit mention of this has been included in the Participant Information Sheets.

### 12. Funding

Funding for the WISE evaluation is provided by the Children’s Investment Fund Foundation (CIFF).

### 13. Ethical considerations

This study will be conducted in accordance with ICH-GCP E6 guidelines. Informed consent to participate in the study will be obtained prior to any study procedure, and necessary steps will be taken to ensure the confidentiality of study participants. Participants or their parents can suspend their participation at any time without any consequence. This study will be subject to LSHTM and national ethical approval in Ethiopia prior to the enrolment of any study participant. The investigators of the study have training in good clinical practice and research ethics.

**Indemnity:** London School of Hygiene & Tropical Medicine holds Public Liability ("negligent harm") and Clinical Trial ("non-negligent harm") insurance policies which apply to this trial.
**Sponsor:** London School of Hygiene & Tropical Medicine will act as the main sponsor for this study. Delegated responsibilities will be assigned locally.

63. UNICEF, *Raising even more clean hands: Advancing health, learning and equity through WASH in schools.* New York, 2012.

64. UNICEF, *WHO. Core Questions and Indicators for Monitoring WASH in Schools in the Sustainable Development Goals.* World Health Organization: Geneva, Switzerland, 2016.

77. UNICEF, *Drinking water, sanitation and hygiene in schools: global baseline report 2018.* 2018.

78. Tamiru, S., et al., *Girls in control: compiled findings from studies on menstrual hygiene management of schoolgirls.* Addis Abbaba, Ethiopia: SNV Netherlands Development Organisation, 2014.

79. Erulkar, A.S., et al. *Ethiopia Young adult SurvEY a StudY in SEvEn rEgionS Population Council*. 2010.

80. MoE (Ministry of Education, E., *Education Statistics Annual Abstract 2011/12*. 2012, MoE Addis Ababa.

81. World Bank, *World Development Report 2012 : Gender Equality and Development.* 2012, World Bank.

82. Abera, Y., *Menarche, Menstruation Related Problems and Practices among Adolescent High School Girls in Addis Ababa, 2003/04.* Addis Ababa University, 2004.

83. Goodman, R., H. Meltzer, and V. Bailey, *The Strengths and Difficulties Questionnaire: a pilot study on the validity of the self-report version.* Int Rev Psychiatry, 2003. **15**(1-2): p. 173-7.

84. Hennegan, J., et al., *Measuring menstrual hygiene experience: development and validation of the Menstrual Practice Needs Scale (MPNS-36) in Soroti, Uganda.* BMJ Open, 2020. **10**(2): p. e034461.

85. Fiske, E.B., *World atlas of gender equality in education*. 2012: Unesco.

86. Drummond, M.F., et al., *Methods for the economic evaluation of health care programmes*. 2015: Oxford university press.

87. Baqui, A.H., et al., *Methodological issues in diarrhoeal diseases epidemiology: definition of diarrhoeal episodes.* Int J Epidemiol, 1991. **20**(4): p. 1057-63.

88. Arnold, B.F., et al., *Optimal recall period for caregiver-reported illness in risk factor and intervention studies: a multicountry study.* Am J Epidemiol, 2013. **177**(4): p. 361-70.

89. StataCorp, *Stata Statistical Software: Release 15*. 2017: College Station, TX: StataCorp LLC.

90. R Core Team, *R: A language and environment for statistical computing.* 2013.

91. Vassall, A., et al., *Reference Case for Estimating the Costs of Global Health Services and Interventions*. 2017.

92. Belachew, T., et al., *Food insecurity, school absenteeism and educational attainment of adolescents in Jimma Zone Southwest Ethiopia: a longitudinal study.* Nutr J, 2011. **10**(1): p. 29.

**Supplementary file B: Project WISE infrastructure and delivery**

Project WISE infrastructure components were delivered to meet *Splash* target ratios (full details of programme standards can be found at https://splash-program-standards.screenstepslive.com):

- 1 drinking tap per 75 users (including adults/staff), based on the single largest shift population at a campus.
- 1 handwashing tap per 75 users (including adults/staff), based on the single largest shift population at a campus.
- A minimum of 9 L/person/day of water storage capacity, based on the total population of users at a school campus. In Addis Ababa, the target is for each school to have 3 days of water storage supply available.

Project WISE intervention schools were provided with water storage tanks to meet storage capacity standards (e.g., Supplementary Figure 1, left) and water filtration systems for to supply drinking water stations (e.g., Supplementary Figure 1, right), meeting the minimum ratio for drinking taps to users (1:75) and the minimum acceptable flow rate per tap (3 L/min). Water quality is tested pre- and post-implementation and routinely throughout the monitoring period.

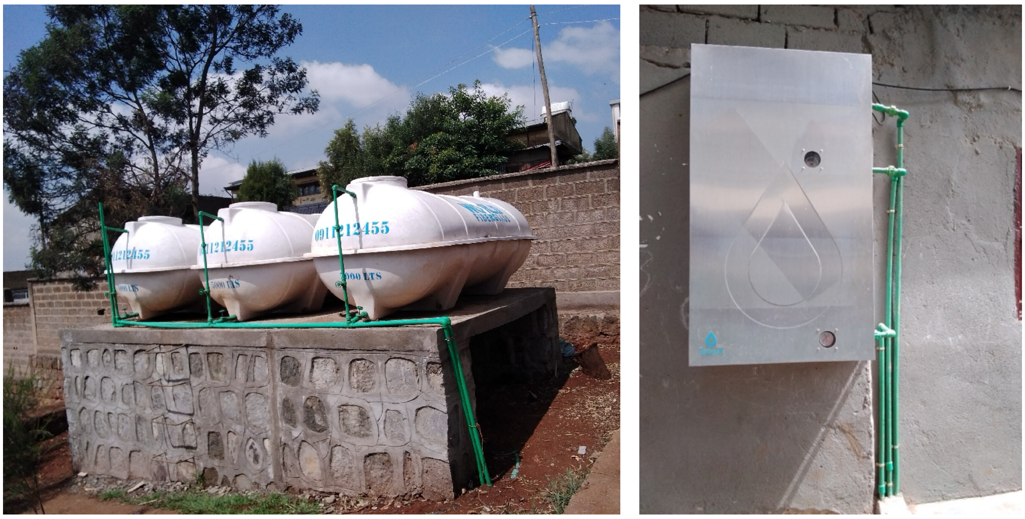

**Supplementary Figure 1. Water storage tanks (left) and water filtration system (right) installed in study schools.**

Drinking water and handwashing stations are made from durable fade-resistant HDPE with recycled material, with different colours and basin depths for handwashing and drinking water to promote separation of behaviours and prevent drinking unfiltered water from handwashing stations. An optional riser at the base is intended to allow the station to serve children of various ages, and stations have smooth surfaces with rounded corners for ease of cleaning.

Drinking water stations (Supplementary Figure 2) had two bottle filler taps per station and a bubbler tap, and were installed in easily visible and accessible locations for the student population, which can include: near play areas, classroom buildings, or the feeding area. Stations were installed on impervious surfaces such as flat stones or concrete, with a concrete pad provided if no suitable surface was available, and with suitable drainage.

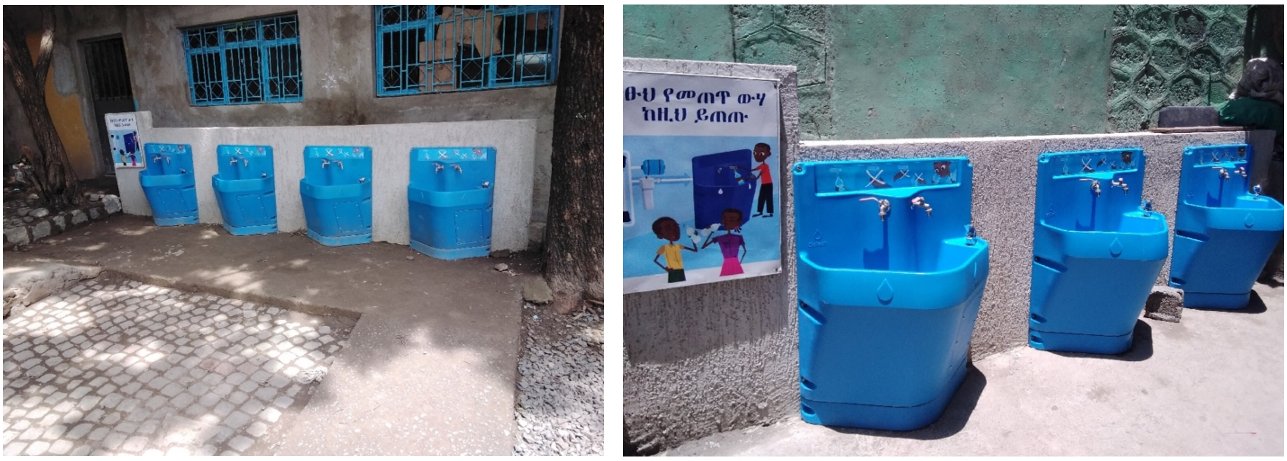

**Supplementary Figure 2. Drinking water stations installed in study schools.**

Handwashing stations (Supplementary Figure 3) had two taps per station, and the sides angled at 45 degrees were designed to promote improved hygiene behaviour through face-to-face handwashing. Handwashing stations were installed to meet targets of at least one handwashing tap for every three toilet stalls and/or urinal spaces, located within 5 metres of the sanitation facility, and at least one handwashing station near the feeding area of the school. Stations were also installed on impervious surfaces such as flat stones or concrete, with a concrete pad provided if no suitable surface was available, and with suitable drainage.

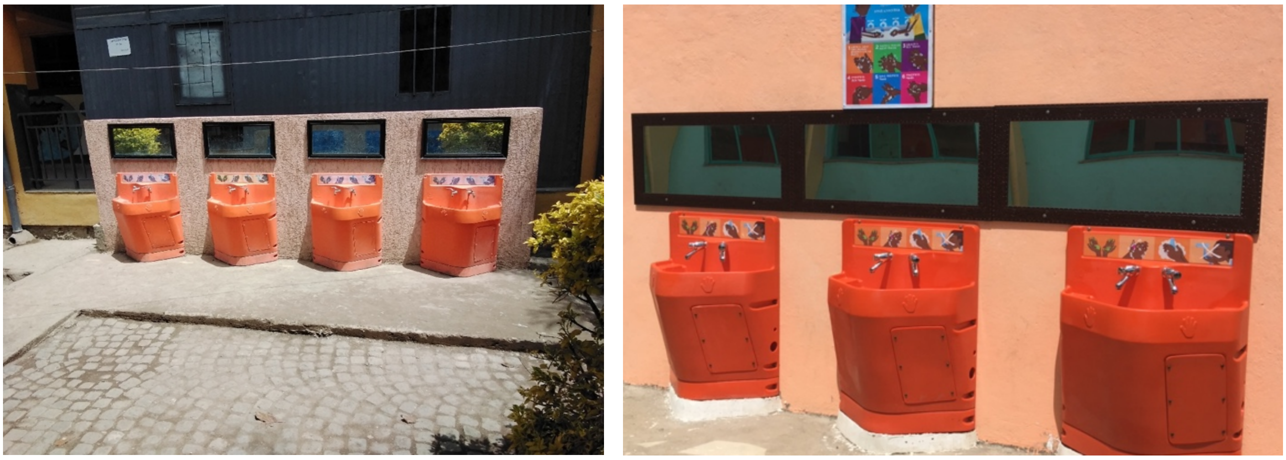

**Supplementary Figure 3. Handwashing stations installed in study schools.**

Mirrors made of a shatter-proof material or with a protective clear cover (acrylic) were installed above every handwashing station (both new and existing stations) at an appropriate angle for children. Posters and signs at both drinking water and handwashing stations (Supplementary Figure 4) provided environmental cues for behaviour and instruction on handwashing technique and correct use of facilities.

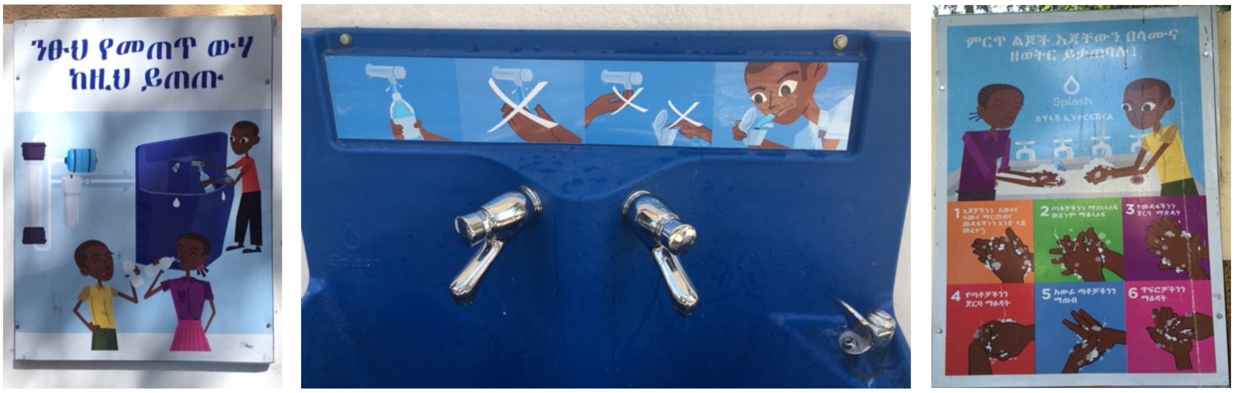

**Supplementary Figure 4. Posters and signage at drinking water stations (left and centre) and handwashing stations (right) installed in study schools.**

The sanitation components of Project WISE (not delivered until after study completion) were planned to meet target ratios of 1 sanitation fixture per 75 male students, and 1 sanitation fixture per 70 female students. Facilities were to be gender-segregated, separate for students and teachers, wheelchair-accessible, well-lit and ventilated, able to close and lock from the inside, with adequate lighting in the stall and hallway, and a water tap and bucket and waste bin for menstrual hygiene management.

The water and hygiene infrastructure improvements (along with core training modules) were delivered to all 30 intervention schools by January 2022. No control school received any intervention component until after study completion. Intervention and control schools were distributed over a wide area across Addis Ababa (Supplementary Figure 5).

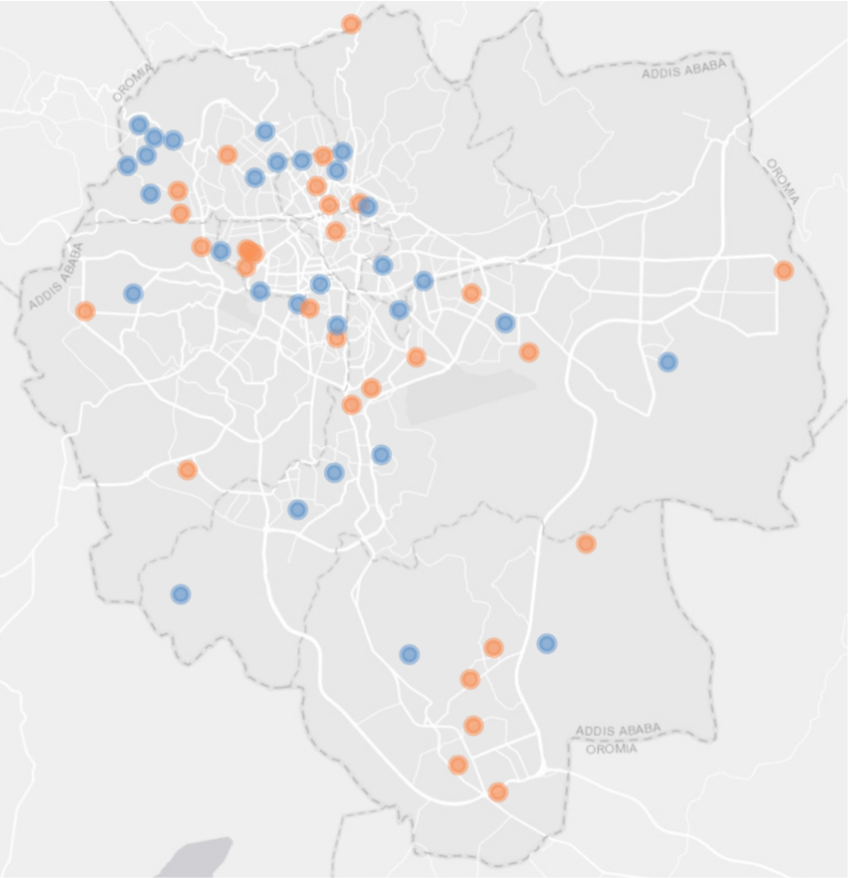

**Supplementary Figure 5. Approximate locations of 30 intervention (blue) and 30 control (orange) schools enrolled in the WISE trial across Addis Ababa, Ethiopia.**

**Supplementary file C: Supplementary tables**

| | Follow-up 1 | | Follow-up 2 | | Follow-up 3 | | Final follow-up | | Total participants | | Total observations | |
|---|---|---|---|---|---|---|---|---|---|---|---|---|
| | Control | Intervention | Control | Intervention | Control | Intervention | Control | Intervention | Control | Intervention | Control | Intervention |
| Roll-call absence | 3088 | 3070 | 3088 | 3072 | 3084 | 3064 | 3054 | 3020 | 3089 | 3079 | 12314 | 12226 |
| Pupil-reported diarrhoea in past 7 days | 2886 | 2861 | 2871 | 2868 | 2600 | 2616 | 2658 | 2573 | 3075 | 3069 | 11015 | 10918 |
| Pupil-reported diarrhoea in past 2 days | 2887 | 2860 | 2870 | 2868 | 2600 | 2616 | 2657 | 2574 | 3075 | 3070 | 11014 | 10918 |
| Pupil-reported respiratory illness in past 7 days | 2882 | 2855 | 2870 | 2865 | 2598 | 2616 | 2653 | 2573 | 3075 | 3070 | 11003 | 10909 |
| Pupil-reported respiratory illness in past 2 days | 2882 | 2857 | 2870 | 2867 | 2601 | 2616 | 2655 | 2574 | 3075 | 3070 | 11008 | 10914 |
| Pupil-reported full-day absence in past week | 2887 | 2862 | 2871 | 2868 | 2602 | 2617 | 2658 | 2575 | 3075 | 3070 | 11018 | 10922 |
| (total days of reporting; approx. 5 in past week) | (14488) | (14341) | (14392) | (14339) | (13010) | (13085) | (13377) | (12952) | | | (55267) | (54717) |
| SDQ-15 total difficulties score | | | | | | | 1829 | 1675 | 1829 | 1675 | 1829 | 1675 |
| SAMNS-26 total score | | | | | | | 545 | 438 | 545 | 438 | 545 | 438 |
| MPNS-36 total score | | | | | | | 530 | 406 | 530 | 406 | 530 | 406 |

**Supplementary Table 1. Number of participants and observations included in assessment of each outcome at each time-point.**

*Note:* Total observations included in assessments at each follow-up do not always exactly match number of eligible participants in Figure 1 due to circumstances where absence could not be assessed (e.g., sentinel classes not available for data collection on the follow-up day), or pupil refusals for specific questions.

| | Primary analysis (adjusted for stratification variables) | | Further adjusted for grade and gender | | Further adjusted for sub-city and time-point* | |
|---|---|---|---|---|---|---|
| | aOR (95% CI) | p-value | aOR (95% CI) | p-value | aOR (95% CI)* | p-value |
| Pupil-reported diarrhoea in past 7 days† | 1·15 (0·83, 1·59) | 0·39 | 1·11 (0·82, 1·52) | 0·50 | 0·95 (0·74, 1·24) | 0·72 |
| Pupil-reported diarrhoea in past 2 days | 1·22 (0·82, 1·83) | 0·32 | 1·19 (0·81, 1·75) | 0·39 | 0·97 (0·71, 1·33) | 0·86 |
| Pupil-reported respiratory illness in past 7 days† | 0·84 (0·71, 1·00) | 0·046 | 0·83 (0·69, 0·98) | 0·031 | 0·83 (0·70, 0·99) | 0·035 |
| Pupil-reported respiratory illness in past 2 days | 0·88 (0·73, 1·06) | 0·18 | 0·87 (0·73, 1·05) | 0·14 | 0·88 (0·73, 1·05) | 0·16 |
| | aOR (95% CI) | p-value | aOR (95% CI) | p-value | aOR (95% CI)* | p-value |
| Roll-call absence† | 1·07 (0·83, 1·38) | 0·59 | 1·08 (0·84, 1·40) | 0·53 | 1·05 (0·81, 1·36) | 0·71 |
| | aOR (95% CI) | p-value | aOR (95% CI) | p-value | aOR (95% CI)* | p-value |
| Pupil-reported full-day absence in past week | 1·01 (0·75, 1·36) | 0·93 | 0·99 (0·74, 1·33) | 0·97 | 1·01 (0·76, 1·36) | 0·94 |
| | aMD (95% CI) | p-value | aMD (95% CI) | p-value | aMD (95% CI) | p-value |
| SDQ-15 total difficulties score (0–40) | 0·03 (-0·62, 0·68) | 0·94 | -0·001 (-0·65, 0·64) | 1·00 | 0·03 (-0·59, 0·65) | 0·94 |
| SAMNS-26 total score (0–100) | 3·32 (0·05, 6·59) | 0·046 | 3·66 (0·29, 7·04) | 0·033 | 3·41 (0·10, 6·72) | 0·043 |
| MPNS-36 total score (0–3) | -0·01 (-0·08, 0·06) | 0·81 | 0·004 (-0·06, 0·07) | 0·91 | 0·01 (-0·04, 0·07) | 0·64 |

**Supplementary Table 2. Intervention effects on primary and secondary outcomes across three models with various adjustments.**

*Note*: Analyses include all eligible children with outcome data at the relevant follow-up. For repeated measures, we calculated a proportion of available follow-ups with illness / absent for each participant, and the mean and SD of these proportions across participants are shown. Stratification variables adjusted for in all models are school size (< or ≥1200 pupils) and presence of a kindergarten. All analyses included a random effect for school, and analyses based on repeated measures included an additional random effect for pupil-level clustering and assumed a constant treatment effect across time-points. *Adjustments for time-point made for repeated measures. †Primary outcomes.

| | Control | | Intervention | | Intervention effect | | |
|---|---|---|---|---|---|---|---|
| | Pupils | Mean prop. of follow-ups with illness (SD) | Pupils | Mean prop. of follow-ups with illness (SD) | aMD (95% CI) | p-value | ICC |
| Pupil-reported diarrhoea in past 7 days† | 3075 | 0·073 (0·151) | 3069 | 0·083 (0·160) | 0·010 (-0·011, 0·031) | 0·35 | 0·062 |
| Pupil-reported diarrhoea in past 2 days | 3075 | 0·042 (0·112) | 3070 | 0·050 (0·127) | 0·009 (-0·006, 0·024) | 0·25 | 0·054 |
| Pupil-reported respiratory illness in past 7 days† | 3075 | 0·276 (0·269) | 3070 | 0·248 (0·257) | -0·032 (-0·062, -0·002) | 0·038 | 0·043 |
| Pupil-reported respiratory illness in past 2 days | 3075 | 0·187 (0·230) | 3070 | 0·171 (0·219) | -0·019 (-0·044, 0·006) | 0·14 | 0·039 |
| | Pupils | Mean prop. of follow-ups absent (SD) | Pupils | Mean prop. of follow-ups absent (SD) | aMD (95% CI) | p-value | ICC |
| Roll-call absence† | 3088 | 0·103 (0·171) | 3078 | 0·106 (0·171) | 0·004 (-0·018, 0·027) | 0·70 | 0·056 |
| | Pupils | Mean prop. of school days reported absent (SD) | Pupils | Mean prop. of school days reported absent (SD) | aMD (95% CI) | p-value | ICC |
| Pupil-reported full-day absence in past week | 3075 | 0·056 (0·090) | 3070 | 0·055 (0·082) | -0·0001 (-0·015, 0·014) | 0·99 | 0·10 |

**Supplementary Table 3. Analysis of mean differences for primary and secondary repeated binary outcomes aggregated at the pupil level.**

*Note*: Analyses include all eligible children with outcome data at the relevant follow-up. For repeated measures, we calculated a proportion of available follow-ups with illness / absent for each participant, and the mean and SD of these proportions across participants are shown. Analysis was based on the mean differences in these pupil-level proportions between trial arms, including a random effect for school. Analyses were adjusted for stratification variables (school size < or ≥1200 pupils, and presence of a kindergarten).
†Primary outcomes.

| | Control | | Intervention | | Intervention effect | | |
|---|---|---|---|---|---|---|---|
| | Pupils | Mean prop. of follow-ups reporting absence (SD) | Pupils | Mean prop. of follow-ups reporting absence (SD) | aOR (95% CI) | p-value | ICC |
| Pupil-reported absence due to illness in past week | 3075 | 0·102 (0·182) | 3069 | 0·093 (0·173) | 0·89 (0·75, 1·05) | 0·17 | 0·017 |
| Pupil-reported absence due to diarrhoea in past week | 3075 | 0·008 (0·047) | 3069 | 0·005 (0·040) | 0·59 (0·37, 0·93) | 0·024 | 0·054 |
| Pupil-reported absence due to respiratory illness in past week | 3075 | 0·042 (0·117) | 3069 | 0·039 (0·111) | 0·90 (0·72, 1·11) | 0·31 | 0·020 |
| | Pupils | Reported illness at final follow-up (%) | Pupils | Reported illness at final follow-up (%) | aOR (95% CI) | p-value | ICC |
| Pupil-reported earache in past 7 days | 2651 | 54 (2·04) | 2571 | 61 (2·37) | 1·17 (0·77, 1·77) | 0·46 | 0·035 |
| Pupil-reported earache in past 2 days | 2656 | 44 (1·66) | 2573 | 48 (1·87) | 1·13 (0·74, 1·71) | 0·57 | 0·001 |
| | Pupils | Median (IQR) | Pupils | Median (IQR) | Proportional aOR (95% CI) | p-value | ICC |
| Subjective wellbeing – smiley faces visual analogue scale (1–5) | 2658 | 4 (3, 5) | 2575 | 4 (3, 5) | 0·92 (0·72, 1·17) | 0·48 | 0·055 |
| | Pupils | Mean score (SD) | Pupils | Mean score (SD) | aMD (95% CI) | p-value | ICC |
| SAMNS-26 subscales (0–100) | | | | | | | |
| MH preparation and maintenance | 549 | 72·4 (17·6) | 443 | 75·2 (18·9) | 2·96 (-0·20, 6·11) | 0·067 | 0·053 |
| Menstrual pain management | 559 | 65·6 (25·9) | 452 | 68·2 (25·0) | 2·33 (-1·29, 5·95) | 0·21 | 0·016 |
| Executing stigmatised tasks | 562 | 59·9 (26·5) | 453 | 62·3 (27·3) | 2·59 (-2·11, 7·29) | 0·28 | 0·056 |
| MPNS-36 subscales (0–3) | | | | | | | |
| Material and home environment needs | 548 | 2·18 (0·54) | 439 | 2·17 (0·57) | -0·01 (-0·08, 0·06) | 0·77 | 0·004 |
| Transport and school environment needs | 556 | 1·66 (0·82) | 434 | 1·78 (0·77) | 0·13 (-0·02, 0·28) | 0·082 | 0·069 |
| Material reliability concerns | 561 | 1·93 (0·86) | 450 | 1·78 (0·95) | -0·13 (-0·27, 0·01) | 0·070 | 0·034 |
| Change and disposal insecurity | 556 | 1·68 (0·60) | 440 | 1·56 (0·65) | -0·10 (-0·22, 0·01) | 0·083 | 0·072 |
| Reuse needs | 522 | 1·69 (1·23) | 418 | 1·95 (1·07) | 0·20 (-0·09, 0·49) | 0·18 | 0·18 |
| Reuse insecurity | 525 | 2·15 (1·01) | 421 | 1·90 (1·07) | -0·23 (-0·45, -0·01) | 0·036 | 0·10 |
| | Schools | Mean score (SD) | Schools | Mean score (SD) | aMD (95% CI) | p-value | |
| Change in gender parity in enrolment (0–2) | 30 | 0·031 (0·088) | 30 | -0·004 (0·058) | -0·04 (-0·07, 0·004) | 0·075 | |

**Supplementary Table 4. Intervention effects on other outcomes.**

*Note*: Analyses include all eligible children with outcome data at the relevant follow-up. For repeated measures, we calculated a proportion of available follow-ups with illness / absent for each participant, and the mean and SD of these proportions across participants are shown. Analyses were adjusted for stratification variables (school size < or ≥1200 pupils, and presence of a kindergarten). Except for gender parity in enrolment (measured at the school level), all analyses included a random effect for school, and analyses based on repeated measures included an additional random effect for pupil-level clustering and assumed a constant treatment effect across time-points.

| | Primary analysis (adjusted for stratification variables) | | Further adjusted for responsibility for household income | | Primary analysis with no age restrictions | |
|---|---|---|---|---|---|---|
| | aOR (95% CI) | p-value | aOR (95% CI) | p-value | aOR (95% CI) | p-value |
| Pupil-reported diarrhoea in past 7 days† | 1·15 (0·83, 1·59) | 0·39 | 1·16 (0·84, 1·60) | 0·37 | 1·16 (0·84, 1·59) | 0·37 |
| Pupil-reported diarrhoea in past 2 days | 1·22 (0·82, 1·83) | 0·32 | 1·23 (0·82, 1·83) | 0·31 | 1·23 (0·83, 1·83) | 0·30 |
| Pupil-reported respiratory illness in past 7 days† | 0·84 (0·71, 1·00) | 0·046 | 0·84 (0·71, 1·00) | 0·044 | 0·84 (0·70, 0·99) | 0·039 |
| Pupil-reported respiratory illness in past 2 days | 0·88 (0·73, 1·06) | 0·18 | 0·88 (0·74, 1·06) | 0·18 | 0·88 (0·73, 1·05) | 0·16 |
| | aOR (95% CI) | p-value | aOR (95% CI) | p-value | aOR (95% CI) | p-value |
| Roll-call absence† | 1·07 (0·83, 1·38) | 0·59 | 1·07 (0·84, 1·38) | 0·58 | 1·04 (0·81, 1·34) | 0·74 |
| | aOR (95% CI) | p-value | aOR (95% CI) | p-value | aOR (95% CI) | p-value |
| Pupil-reported full-day absence in past week | 1·01 (0·75, 1·36) | 0·93 | 1·01 (0·76, 1·36) | 0·92 | 1·00 (0·75, 1·34) | 0·99 |
| | aMD (95% CI) | p-value | aMD (95% CI) | p-value | aMD (95% CI) | p-value |
| SDQ-15 total difficulties score (0–40) | 0·03 (-0·62, 0·68) | 0·94 | 0·03 (-0·62, 0·68) | 0·93 | 0·05 (-0·61, 0·71) | 0·88 |
| SAMNS-26 total score (0–100) | 3·32 (0·05, 6·59) | 0·046 | 3·34 (0·07, 6·61) | 0·045 | 3·17 (0·006, 6·34) | 0·050 |
| MPNS-36 total score (0–3) | -0·01 (-0·08, 0·06) | 0·81 | -0·01 (-0·08, 0·06) | 0·75 | -0·003 (-0·07, 0·07) | 0·94 |

**Supplementary Table 5. Sensitivity analyses.**

*Note*: Analyses include all eligible children with outcome data at the relevant follow-up. Stratification variables adjusted for in all models are school size (< or ≥1200 pupils) and presence of a kindergarten. All analyses included a random effect for school, and analyses based on repeated measures included an additional random effect for pupil-level clustering and assumed a constant treatment effect across time-points. †Primary outcomes.
